# Total predicted MHC-I epitope load is inversely associated with population mortality from SARS-CoV-2

**DOI:** 10.1101/2020.05.08.20095430

**Authors:** Eric A. Wilson, Gabrielle Hirneise, Abhishek Singharoy, Karen S. Anderson

## Abstract

Polymorphisms in MHC-I protein sequences across human populations significantly impacts viral peptide binding capacity and thus alters T cell immunity to infection. Consequently, allelic variants of the MHC-I protein have been found to be associated with patient outcome to various viral infections, including SARS-CoV. In the present study, we assess the relationship between observed SARS-CoV-2 population mortality and the predicted viral binding capacities of 52 common MHC-I alleles. Potential SARS-CoV-2 MHC-I peptides were identified using a consensus MHC-I binding and presentation prediction algorithm, called EnsembleMHC. Starting with nearly 3.5 million candidates, we resolved a few hundred highly probable MHC-I peptides. By weighing individual MHC allele-specific SARS-CoV-2 binding capacity with population frequency in 23 countries, we discover a strong inverse correlation between the predicted population SARS-CoV-2 peptide binding capacity and observed mortality rate. Our computations reveal that peptides derived from the structural proteins of the virus produces a stronger association with observed mortality rate, highlighting the importance of S, N, M, E proteins in driving productive immune responses. The correlation between epitope binding capacity and population mortality risk remains robust across a range of socioeconomic and epidemiological factors. A combination of binding capacity, number of deaths due to COPD complications, gender demographics. and the proportions of the population that were over the age of 65 and overweight offered the strongest determinant of at-risk populations. These results bring to light how molecular changes in the MHC-I proteins may affect population-level outcomes of viral infection.

## Introduction

In December 2019, the novel coronavirus, SARS-CoV-2 was identified from a cluster of cases of pneumonia in Wuhan, China^1,2^. With over 73.1 million cases and over 1.6 million deaths, the viral spread has been declared a global pandemic by the World Health Organization^3^. Due to its high rate of transmission and unpredictable severity, there is an immediate need for information surrounding the adaptive immune response towards SARS-CoV-2.

A robust T cell response is integral for the clearance of coronaviruses, and generation of lasting immunity^4^. The potential role of T cells for coronavirus clearance has been supported by the identification of immunogenic CD8^+^ T cell epitopes in the S (Spike), N (Nucleocapsid), M (Membrane), and E (Envelope) proteins^5^. Additionally, SARS-CoV specific CD8^+^ T cells have been shown to provide long lasting immunity with memory CD8^+^ T cells being detected up to 17 years post infection^4,6,7^. The specifics of the T cell response to SARS-CoV-2 is still evolving. However, a recent screening of SARS-CoV-2 peptides revealed a majority of the CD8^+^ T cell immune response is targeted towards viral structural proteins (N, M, S)^8^.

A successful CD8^+^ T cell response is contingent on the efficient presentation of viral protein fragments by Major Histocompatibility Complex I (MHC-I) proteins. MHC-I molecules bind and present peptides derived from endogenous proteins on the cell surface for CD8^+^ T cell interrogation. The MHC-I protein is highly polymorphic, with amino acid substitutions within the peptide binding groove drastically altering the composition of presented peptides. Consequently, the influence of MHC genotype to shape patient outcome has been well studied in the context of viral infections^9^. For coronaviruses, there have been several studies of MHC association with disease susceptibility. A study of a Taiwanese and Hong Kong cohort of patients with SARS-CoV found that the MHC-I alleles HLA-B* 07:03 and HLA-B* 46:01 were linked to increased susceptibility while HLA-Cw* 15:02 was linked to increased resistance^10–12^. However, some of the reported associations did not remain after statistical correction, and it is still unclear if MHC-outcome associations reported for SARS-CoV are applicable to SARS-CoV-2^13,14^. Recently, a comprehensive prediction of SARS-CoV-2 MHC-I peptides indicated a relative depletion of high affinity binding peptides for HLA-B* 46:01, hinting at a similar association profile in SARS-CoV-2^15^. More importantly, it remains elusive if such a depletion of putative high affinity peptides will impact patient outcome to SARS-CoV-2 infections.

The lack of large scale genomic data linking individual MHC genotype and outcome from SARS-CoV-2 infections precludes a similar analysis as performed for SARS-CoV^10–12^. Therefore, we endeavored to assess the relationship between the predicted SARS-CoV-2 binding capacity of a population and the observed SARS-CoV-2 mortality rate. However, current MHC-I prediction algorithms have been characterized by a high false positive rate particularly when predicting peptides that are naturally presented^16,17^. To mitigate false positives and identify the highest confidence SARS-CoV-2 MHC-I peptides, we developed a consensus prediction algorithm, coined EnsembleMHC, and predicted MHC-I peptides for a panel of 52 common MHC-I alleles^18^. This prediction workflow integrates seven different algorithms that have been parameterized on high-quality mass spectrometry data and provides a confidence level for each identified peptide^17,19^. The distribution of the number of high-confidence peptides assigned to each allele was used to assess a country-specific SARS-CoV-2 binding capacity, called the EnsembleMHC population score, for 23 countries (for selection criteria, please refer to the methods). This score was derived by weighing the individual binding capacities of the 52 MHC-I alleles by their endemic frequencies. We observe a strong inverse correlation between the EnsembleMHC population score and observed population SARS-CoV-2 mortality. Furthermore, the correlation is shown to become stronger when considering EnsembleMHC population scores based solely on SARS-CoV-2 structural proteins, underlining their potential importance in driving a robust immune response. Based on their predicted binding affinity, expression, and sequence conservation in viral isolates, we identified 108 peptides derived from SARS-CoV-2 structural proteins that are high-value targets for CD8^+^ T cell vaccine development.

## Results

### EnsembleMHC workflow offers more precise MHC-I presentation predictions than individual algorithms

The accurate assessment of differences in SARS-CoV-2 binding capacities across MHC-I allelic variants requires the isolation of MHC-I peptides with a high probability of being presented. EnsembleMHC provides the requisite precision through the use of allele and algorithm-specific score thresholds and peptide confidence assignment.

MHC-I alleles substantially vary in both peptide binding repertoire size and median binding affinity^25^. The EnsembleMHC workflow addresses this inter-allele variation by identifying peptides based on MHC allele and algorithm-specific binding affinity thresholds. These thresholds were set by benchmarking each of the seven component algorithms against 52 single MHC allele peptide data sets^17^. Each data set consists of mass spectrometry-confirmed MHC-I peptides that have been naturally presented by a model cell line expressing one of the 52 select MHC-I alleles. These experimentally validated peptides, denoted target peptides, were supplemented with a 100-fold excess of decoy peptides. Decoys were generated by randomly sampling peptides that were not detected by mass spectrometry, but were derived from the same protein sources as a detected target peptide. Algorithm and allele-specific binding affinity thresholds were then identified through the independent application of each component algorithm to all MHC allele data sets.

For every data set and algorithm combination, the target and decoy peptides were ranked by predicted binding affinity to the MHC allele defined by that data s et. Then, an algorithm-specific binding affinity threshold was set to the minimum score needed to isolated the highest affinity peptides commensurate to 50% of the observed allele repertoire size **(methods, SI A.1)**. The observed allele repertoire size was defined as the total number of target peptides within a given single MHC allele data set. Therefore, if a data set had 1000 target peptides, the top 500 highest affinity peptides would be selected, and the algorithm-specific threshold would be set to the predicted binding affinity of the 500^*th*^ peptide. This parameterization method resulted in the generation of a customized set of allele and algorithm-specific binding affinity thresholds in which an expected quantity of peptides can be recovered.

Consensus MHC-I prediction typically require a method for combining outputs from each individual component algorithm into a composite score. This composite score is then used for peptide selection. EnsembleMHC identifies high-confidence peptides based on filtering by a quantity called *peptide*^*FDR*^ **(methods Eq. 1)**. During the identification of allele and algorithm-specific binding affinity thresholds, the empirical false detection rate (*FDR*) of each algorithm was calculated. This calculation was based on the proportion of target to decoy peptides isolated by the algorithm specific binding affinity threshold. A *peptide*^*FDR*^ is then assigned to each individual peptide by taking the product of the empirical *FDR*s of each algorithm that identified that peptide for the same MHC-I allele. Analysis of the parameterization process revealed that the overall performance of each included algorithms was comparable, and there was diversity in individual peptide calls by each algorithm, supporting an integrated approach to peptide confidence assessment **(SI A.2)**. Peptide identification by EnsembleMHC was performed by selecting all peptides with a *peptide*^*FDR*^ of less than or equal to 5%^26^.

The efficacy of *peptide*^*FDR*^ as a filtering metric was determined through the prediction of naturally presented MHC-I peptides derived from ten tumor samples^17^ **(Figure 1)**. Similar to the single MHC allele data sets, each tumor sample data set consisted of mass spectrometry-detected target peptides and a 100-fold excess of decoy peptides. The relative performance of EnsembleMHC was assessed via comparison with individual component algorithms. Peptide identification by each algorithm was based on a restrictive or permissive binding affinity thresholds **(Figure 1A (inset table))**. For the component algorithms, the permissive and restrictive thresholds correspond to commonly used binding affinity cutoffs for the identification of weak and strong binders, respectively^27^. The performance of each algorithm on the ten data sets was evaluated through the calculation of the empirical precision, recall, and F1 score.

**Figure 1.**
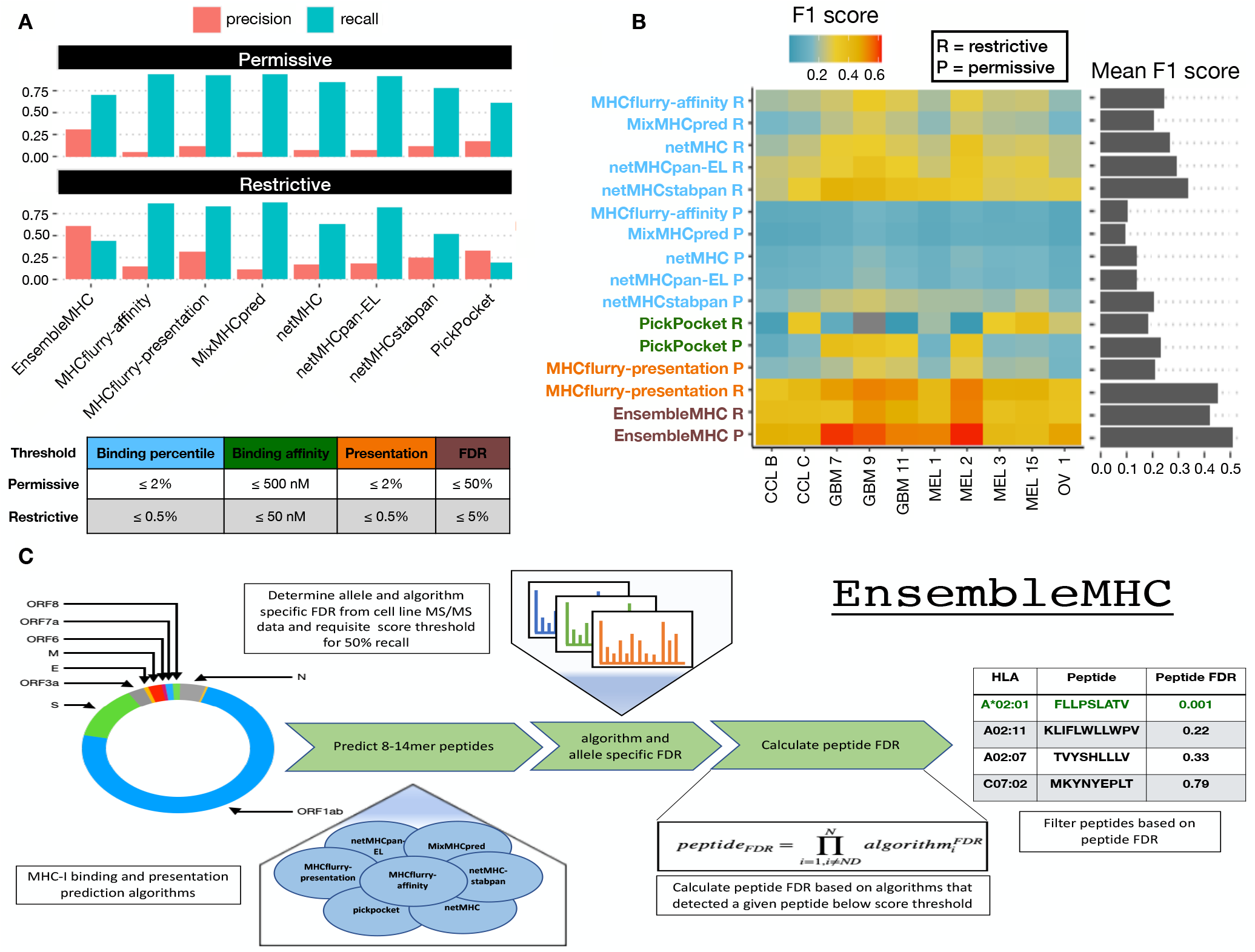
Application of the EnsembleMHC prediction algorithm. The EnsembleMHC prediction algorithm was used to recover MHC-I peptides from 10 tumor sample data sets. **A,** The average precision and recall for EnsembleMHC and each component algorithm was calculated across all 10 tumor samples. Peptide identification by each algorithm was based on commonly used restrictive (strong) or permissive (strong and weak) binding affinity thresholds **(inset table). B,** The F1 score of each algorithm was calculated for all tumor samples. Each algorithm is grouped into 1 of 4 categories: binding affinity represented by percentile score (blue), binding affinity represented by predicted peptide IC50 value (green), MHC-I presentation prediction (orange), and EnsembleMHC (brown). The heatmap colors indicate the value of the observed F1 score (color bar) for a given algorithm (y-axis) on a particular data set (x-axis). Warmer colors indicate higher F1 scores, and cooler colors indicate lower F1 scores. The average F1 score for each algorithm across all samples is shown in the marginal bar plot. **C,** The schematic for the application of the EnsembleMHC predication algorithm to identify SARS-CoV-2 MHC-I peptides.

The average precision and recall of each algorithm across all tumor samples demonstrated an inverse relationship **(Figure 1A)**. In general, restrictive binding affinity thresholds produced higher precision at the cost of poorer recall. When comparing the precision of each algorithm at restrictive thresholds, EnsembleMHC demonstrated a 3.4-fold improvement over the median precision of individual component algorithms. EnsembleMHC also produced the highest F1 score with an average of 0.51 followed by mhcflurry-presentation with an F1 score of 0.45, both of which are 1.5-2 fold higher than the rest of the algorithms **(Figure 1B)**. result was shown to be robust across a range of *peptide*^*FDR*^ cutoff thresholds **(SI A.3)** and alternative performance metrics **(SI A.4)**. Furthermore, EnsembleMHC demonstrated the ability to more efficiently prioritize peptides with experimentally established immunogenicity from the Hepatitis-C genome polyprotein, the Dengue virus genome polyprotein, and the HIV-1 POL-GAG protein **(SI A.5)**. Taken together, these results demonstrate the enhanced precision of EnsembleMHC over individual component algorithms when using common binding affinity thresholds.

In summary, the EnsembleMHC workflow offers two desirable features. First, it determines allele-specific binding affinity thresholds for each algorithm at which a known quantity of peptides are expected to be successfully presented on the cell surface. Second, it assigns a confidence level to each peptide call made by each algorithm. Together, these traits enhance the ability to identify MHC-I peptides with a high probability of successful cell surface presentation.

EnsembleMHC was used to identify MHC-I peptides for the SARS-CoV-2 virus **(Figure 1C)**. The resulting identification of high-confidence SARS-CoV-2 peptides allows for the characterization of alleles that are enriched or depleted for predicted MHC-I peptides. The resulting distribution of allele-specific SARS-CoV-2 binding capacities will then be weighed by the normalized frequencies of the 52 alleles **(SI A.6, Methods Eq. 5-6)** in 23 countries to determine the population-specific SARS-CoV-2 binding capacity or EnsembleMHC population score **(Methods Eq. 7)**. The potential impact of varying population SARS-CoV-2 binding capacities on disease outcome can then be assessed by correlating population SARS-CoV-2 mortality rates with EnsembleMHC population scores. Below, we use EnsembleMHC population scores to stratify countries based on their mortality risks.

### The MHC-I peptide-allele distribution for SARS-CoV-2 structural proteins is especially disproportionate

MHC-I peptides derived from the SARS-CoV-2 proteome were predicted and prioritized using EnsembleMHC. A total of 67,207 potential 8-14mer viral peptides were evaluated for each of the considered MHC-I alleles. After filtering the pool of candidate peptides at the 5% *peptide*^*FDR*^ threshold, the number of potential peptides was reduced from 3.49 Million to 971 (658 unique peptides) **(SI A.7, SI table B.1)**. Illustrated in **Figure 2A**, the viral peptide-MHC allele (or peptide-allele) distribution for high-confidence SARS-CoV-2 peptides was determined by assigning the identified peptides to their predicted MHC-I alleles. There was a median of 16 peptides per allele with a maximum of 47 peptides (HLA-A* 24:02), a minimum of 3 peptides (HLA-A* 02:05), and an interquartile range (IQR) of 16 peptides. Quality assurance of the predicted peptides was performed by computing the peptide length frequencies and binding motifs. The predicted peptides were found to adhere to expected MHC-I peptide lengths^28^ with 78% of the peptides being 9 amino acids in length, 13% being 10 amino acids in length, and 8% of peptides accounting for the remaining lengths **(SI A.8)**. Similarly, logo plots generated from predicted peptides were found to closely reflect reference peptide binding motifs for considered alleles^29^**(SI A.9)**. Overall, the EnsembleMHC prediction platform demonstrated the ability to isolate a short list of potential peptides which adhere to expected MHC-I peptide characteristics.

**Figure 2.**
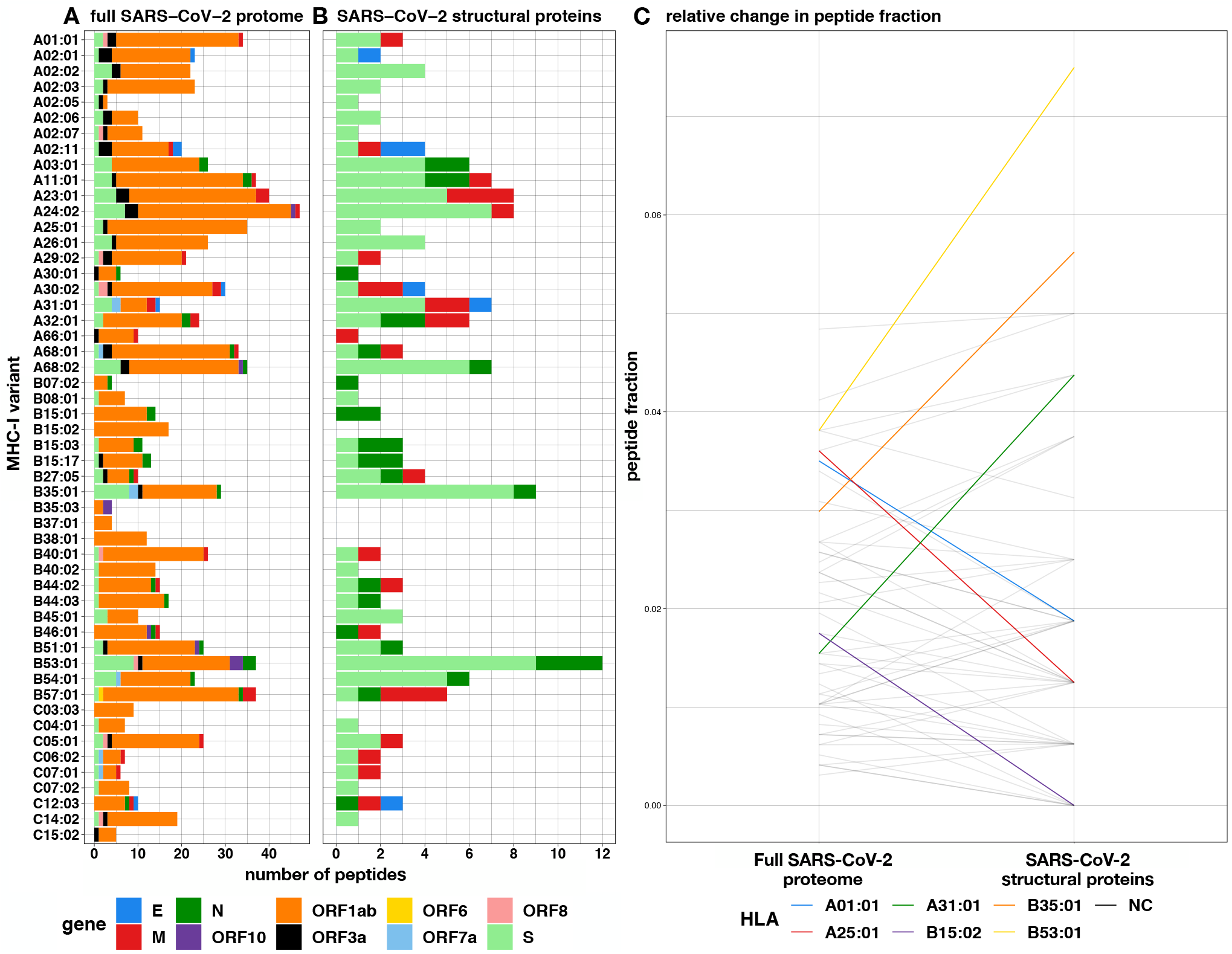
Prediction of SARS-CoV-2 peptides across 52 common MHC-I alleles. **A-B,** The EnsembleMHC workflow was used to predict MHC-I peptides for 52 alleles from the entire SARS-CoV-2 proteome or specifically SARS-CoV-2 structural proteins (envelope, spike, nucleocapsid, and membrane). **C,** The peptide fractions for both protein sets were calculated by dividing the number of peptides assigned to a given allele by the total number of identified peptides for that protein set. Each line indicates the change in peptide fraction observed by a given allele when comparing the viral peptide-MHC allele distribution for the full SARS-CoV-2 proteome proteome or structural proteins. Alleles showing a change of greater than the median peptide fraction, 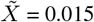, are highlighted in color. For the performance of EnsmebleMHC at a range of different *peptide*^*F DR*^ cutoff thresholds, refer to **SI A.3**

The high expression, relative conservation, and reduced search space of SARS-CoV-2 structural proteins (S, E, M, and N) makes MHC-I binding peptides derived from these proteins high-value targets for CD8^+^ T cell-based vaccine development. **Figure 2B** describes the peptide-allele distribution for predicted MHC-I peptides originating from the four structural proteins. This analysis markedly reduces the number of considered peptides from 658 to 108 **(SI table B.1)**. The median number of predicted SARS-CoV-2 structural peptides assigned to each MHC-I allele was found to be 2 with a maximum of 12 peptides (HLA-B* 53:01), a minimum of 0 (HLA-B* 15:02, B* 35:03,B* 38:01,C* 03:03,C* 15:02), and a IQR of 3 peptides. Analysis of the molecular source of the identified SARS-CoV-2 structural protein peptides revealed that they originate from enriched regions that are highly conserved **(SI A.10-A.11)**. This indicates that such peptides would be good candidates for targeted therapies as they are unlikely to be disrupted by mutation, and several peptides can be targeted using minimal stretches of the source protein. Altogether, consideration of MHC-I peptides derived only from SARS-CoV-2 structural proteins reduces the number of potential peptides to a condensed set of high-value targets that is amenable to experimental validation.

Both the peptide-allele distributions, namely the ones derived from the full SARS-CoV-2 proteome and those from the structural proteins, were found to significantly deviate from an even distribution of predicted peptides as apparent in **figure 2AB** and reflected in the Kolmogorov–Smirnov test p-values (**SI A.12**, full proteome = 5.673e-07 and structural proteins = 1.45e-02). These results support a potential allele-specific hierarchy for SARS-CoV-2 peptide presentation.

To determine if the MHC-I binding capacity hierarchy was consistent between the full SARS-CoV-2 proteome and SARS-CoV-2 structural proteins, the relative changes in the observed peptide fraction (number of peptides assigned to an allele / total number of peptides) between the two protein sets was visualized **(Figure 2C)**. Six alleles demonstrated changes greater than the median peptide fraction 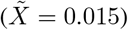 when comparing the two protein sets. The greatest decrease in peptide fraction was observed for A* 25:01 (1.52 times the median peptide fraction), and the greatest increase was seen with B* 53:01 (2.38 times the median peptide fraction). Furthermore, the resulting SARS-CoV-2 structural protein peptide-allele distribution was found to be more variable than the distribution derived from the full SARS-CoV-2 proteome with a quartile coefficient of dispersion of 0.6 compared to 0.44, respectively. This indicates that peptides derived from SARS-CoV-2 structural proteins experience larger relative inter-allele binding capacity discrepancies than peptides derived from the the full SARS-CoV-2 proteome. Together, these results indicate a potential MHC-I binding capacity hierarchy that is more pronounced for SARS-CoV-2 structural proteins.

### Total population epitope load inversely correlates with reported death rates from SARS-CoV-2

The documented importance of MHC-I peptides derived from SARS-CoV-2 structural proteins^8^, coupled with the observed MHC allele binding capacity hierarchy and the high immunogenicity rate of SARS-CoV-2 structural protein MHC-I peptides identified by EnsembleMHC (95% peptides tested *in vitro*, SI A.13), prompts a potential relationship between MHC-I genotype and infection outcome. However, due to the absence of MHC genotype data for SARS-CoV-2 patients, we assessed this relationship at the population-level by correlating predicted country-specific SARS-CoV-2 binding capacity (or EnsembleMHC population score) with observed SARS-CoV-2 mortality.

EnsembleMHC population scores (EMP) were determined for 23 countries **(SI B.2)** by weighing the individual binding capacities of 52 common MHC-I alleles by their normalized endemic expression^18^ **(methods, SI A.6)**. This results in every country being assigned two separate EMP scores, one calculated with respect to the 108 unique SARS-CoV-2 structural protein peptides (structural protein EMP) and the other with respect to the 658 unique peptides derived from the full SARS-CoV-2 proteome (full proteome EMP). The EMP score corresponds to the average predicted SARS-CoV-2 binding capacity of a population. Therefore, individuals in a country with a high EMP score would be expected, on average, to present more SARS-CoV-2 peptides to CD8^+^ T cells than individuals from a country with a low EMP score. The resulting EMP scores were then correlated with observed SARS-CoV-2 mortality (deaths per million) as a function of time. Temporal variance in community spread within the cohort of countries was corrected by truncating the SARS-CoV-2 mortality data set for each country to start after a certain minimum death threshold was met. For example, if the minimum death threshold was 50, then day 0 would be when each country reported at least 50 deaths. The number of countries included in each correlation decreases as the number of days increases due to discrepancies in the length of time that each country met a given minimum death threshold **(SI table B.3)**. Therefore, the correlation between EMP score and SARS-CoV-2 mortality was only estimated at time points where there were at least eight countries. The eight country threshold was chosen because it is the minimum sample size needed to maintain sufficient power when detecting large effect sizes (*ρ* > 0.85). The strength of the relationship between EMP score and SARS-CoV-2 mortality was determined using Spearman’s rank-order correlation (for details concerning the choice of statistical tests, please refer to the methods section). Accordingly, both EMP scores and SARS-CoV-2 mortality data were converted into ascending ranks with the lowest rank indicating the minimum value and the highest rank indicating the maximum value. For instance, a country with an EMP score rank of 1 and death per million rank of 23 would have the lowest predicted SARS-CoV-2 binding capacity and the highest level of SARS-CoV-2-related mortality. Using the described paradigm, the structural protein EMP score and the full proteome EMP score were correlated with SARS-CoV-2-related deaths per million for 23 countries.

Total predicted population SARS-CoV-2 binding capacity exhibited a strong inverse correlation with observed deaths per million. This relationship was found to be true for correlations based on the structural protein EMP **(Figure 3A)** and full proteome EMP **(SI A.14)** scores with a mean effect size of −0.66 and −0.60, respectively. Significance testing of the correlations produced by both EMP scores revealed that the majority of reported correlations are statistically significant with 63% attaining a p-value of ≤ 0.05. Correlations based on the structural protein EMP score demonstrated a 24% higher proportion of statistically significant correlations compared to the full proteome EMP score (74% vs 51%). Furthermore, correlations for EMP scores based on structural proteins produced narrower 95% confidence intervals **(SI A.15-A.16**, **SI table B.3)**. Due to relatively low statistical power of the obtained correlations **(SI A.17)**, the positive predictive value for each correlation **(methods, Eq. 8)** was calculated. The resulting proportions of correlations with a positive predictive value of ≥ 95% were similar to the observed significant p-value proportions with 62% of all measured correlations, 72% of structural protein EMP score correlations, and 52% full proteome EMP score correlations **(SI A.14)**. The similar proportions of significant p-values and PPVs supports that an overall true association is being captured. Furthermore, analysis of similar sized peptide sets sampled from the full SARS-CoV-2 proteome revealed that the observed distinction between the correlations produced by the two protein groups are unlikely to be due to differences in peptide set sizes **(SI A.18)**

**Figure 3.**
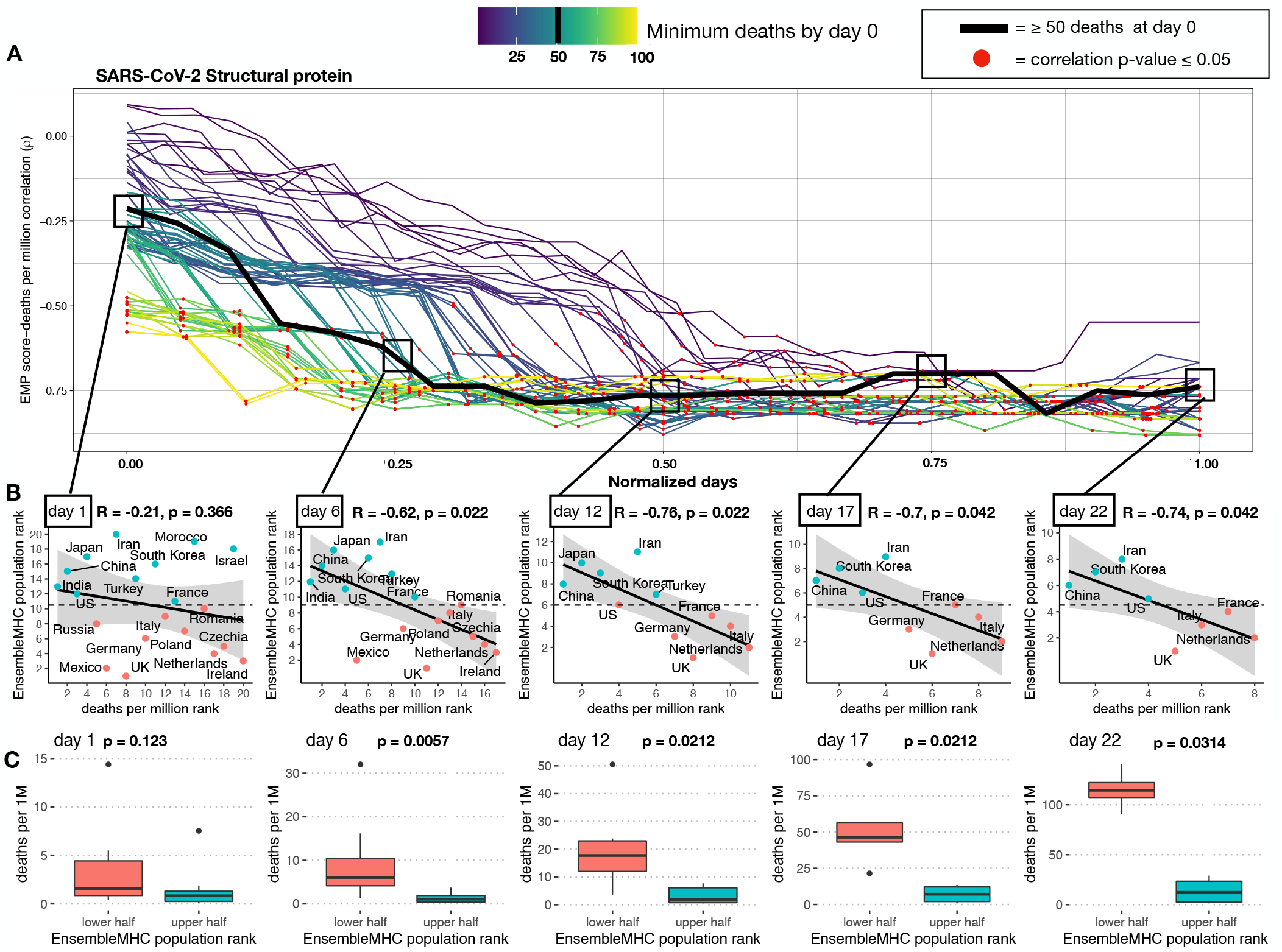
Predicted total epitope load within a population inversely correlates with mortality. **A,** SARS-CoV-2 structural protein-based EnsembleMHC population scores were assigned to 23 countries **(SI B.2),** and correlated with observed mortality rate (deaths per million). The correlation coefficient is presented as a function of time. Individual country mortality rate data were aligned by truncating each data set to start after a minimum threshold of deaths was observed in a given country **(line color).** The Spearman’s rank correlation coefficient between structural protein EMP score and SARS-CoV-2 mortality rate was calculated at every day following day 0 for each of the minimum death thresholds. Due to the differing lengths of time series analysis at each minimum death threshold, the number of days were normalized to improve visualization. Thus, normalized day 0 represents the day when qualifying countries recorded at least the number of deaths indicated by the minimum death threshold, and normalized day 1 represents the final time point at which a correlation was measured. (For mapping between real and normalized days, see **SI B.2**). Correlations that were shown to be statistically significant (p-value ≤ 0.05) are indicated by a red point. **B**, The correlations between the structural protein EnsembleMHC population score (y-axis) and deaths per million (x-axis) were shown for countries meeting the 50 minimum deaths threshold at days 1, 6, 12, 17, and 22. Correlation coefficients and p-values were assigned using Spearman’s rank correlation and the shaded region signifies the 95% confidence interval. Due to Spearman’s rank correlation only considering data rank, Deaths per million and EnsembleMHC population score were converted to ascending rank values (low rank = low values, high rank = high values) to improve visualization of the measured relationship. Red points indicate a country that has an EnsembleMHC population rank less than the median EnsembleMHC population rank of all countries at that day, and blue points indicate a country with an EnsembleMHC population rank greater than the median EnsembleMHC population rank. **C**, The countries at each day were partitioned into a upper or lower half based on the median observed EnsembleMHC population rank. Therefore, countries with an EnsembleMHC population rank greater than the median group EnsembleMHC population score were assigned to the upper half (red), and the remaining countries were assigned to the lower half (blue). p-values were determined by Mann-Whitney U test. The presented box plots are in the style of Tukey (box defined by 25%, 50%, 75% quantiles, and whiskers *±* 1.5 × IQR). The increasing gap between the red and the blue box plots indicates a greater discrepancy in the number of deaths per million between the two groups. The p values in all figures were corrected using the Benjamini-Hochberg procedure^30^ relative to the number of tests performed for each death threshold

Finally, the reported correlations did not remain after randomizing the allele assignment of predicted peptides prior to *peptide*^*FDR*^ filtering **(SI A.19)**, through the use of any individual algorithm **(SI A.20)**. This indicates that the observed relationship is contingent on the high-confidence peptide-allele distribution produced by the EnsembleMHC prediction algorithm. Altogether, these data demonstrate that the MHC-I allele hierarchy characterized by EnsembleMHC is inversely associated with SARS-CoV-2 population mortality, and that the relationship becomes stronger when considering only the presentation of SARS-CoV-2 structural proteins.

The ability to use structural protein EMP score to identify high and low risk populations was assessed using the median minimum death threshold (50 deaths) at evenly spaced time points **(Figure 3A**, **squares)**. All correlations, with the exception of day 1, were found to be significant with an average effect size of −0.71 **(Figure 3B)**. Next, the countries at each day were partitioned into a high or low group based based on whether their assigned EMP score was higher or lower than the median observed EMP score **(Figure 3C)**. The resulting grouping demonstrated a statistically significant difference in the median deaths per million between countries with low structural protein EMP score and countries with high structural protein EMP scores. Additionally, it was observed that deaths per million increased much more rapidly in countries with low structural protein EMP scores. Taken together, these results indicate that structural protein EMP score may be useful for assessing population risk from SARS-CoV-2 infections.

In summary, we make several important observations. First, there is a strong inverse correlation between predicted population SARS-CoV-2 binding capacity and observed deaths per million. This finding suggests that outcome to SARS-CoV-2 may be tied to total epitope load. Second, the correlation between predicted epitope load and population mortality is stronger for SARS-CoV-2 structural MHC-I peptides. This suggests that CD8^+^ T cell-mediated immune response maybe primarily driven by recognition of epitopes derived from these proteins, a finding supported by recent T cell epitope mapping of SARS-CoV-2^8^. Finally, the EnsembleMHC population score can separate countries within the considered cohort into high or low risk populations.

### Structural protein EMP score correlates better with population outcome than identified individual risk factors

Recent large scale patient studies have identified several socioeconomic and health-related factors associated with increased risk of death from SARS-CoV-2 infections^31,32^. To delineate the relative importance of the structural protein EMP score as a SARS-CoV-2 severity descriptor, 12 additional risk factors were assessed for their ability to model population level SARS-CoV-2 outcome in 21 countries (**SI B.4**).

Overall, the structural protein EMP scores produced a significantly stronger association with population SARS-CoV-2 mortality compared to other 12 descriptors **(Figure 4A)**. While various effect size trends were observed, all additional covariates failed to produce statistically significant correlations. To determine if the modeling of SARS-CoV-2 mortality rate could be improved by the combination of single socioeconomic or health-related risk factors with structural protein EMP scores, a set of linear models consisting of either a single risk factor (single feature model) or that factor combined with structural protein EMP scores (combination model) were generated for every time point across each minimum death threshold **(methods)**. Following model generation, the adjusted coefficient of determination (*R*^2^) and significance level of each individual model was extracted and aggregated by dependent variable **(SI A.21)**. Single feature models were characterized by low 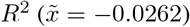 while combination models showed significant improvement 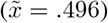. Similarly, combination models demonstrated a substantially higher proportion of statistical significance **(SI A.21B)**. To determine the set of features that produce the best fitting model, all possible combinations of explanatory factors (risk factors and structural protein EMP score) were tested. Subsequently, the top ten performing models, ranked by adjusted *R*^2^ value, were selected for analysis **(Figure 4B)**. The identified models were found to be largely significant (average proportion of significant regressions = 72%) and produce strong fits to the data (average *R*^2^ = 0.7).

**Figure 4.**
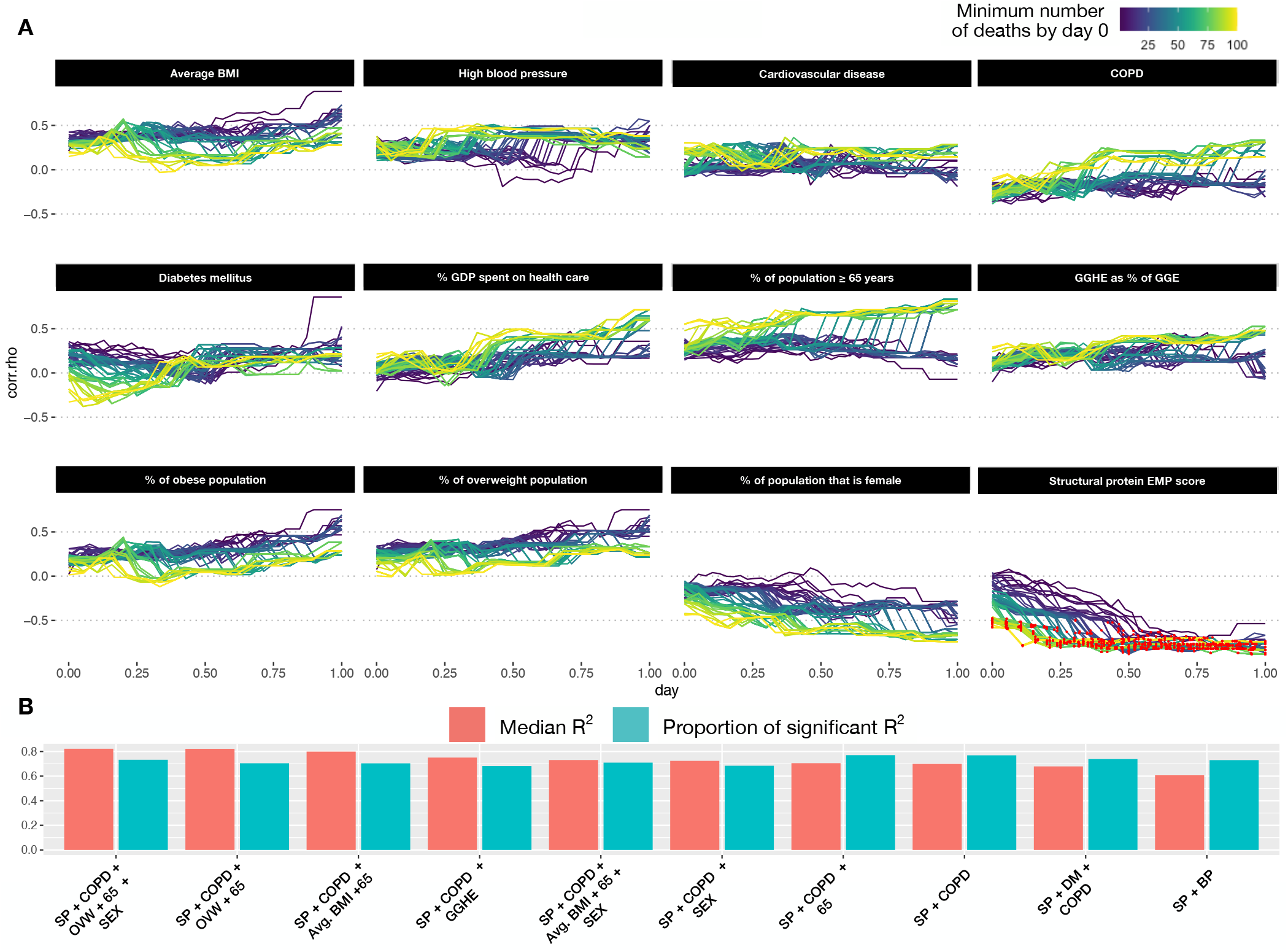
Analysis of other SARS-CoV-2 covariates with observed SARS-CoV-2 population mortality and development of an integrative model. **A,** 12 covariates associated with SARS-CoV-2 mortality on the individual patent level were assessed for correlation with population level mortality **(SI table B.4)**. The correlation of each country-level covariate was determined at each time point after a minimum death threshold was met (line color). The x-axis represents the number of days (normalized) following when a minimum death threshold was met, and the y-axis indicates the observed effect size for that covariate at a given time point. Correlations achieving statistical significance are colored with a red dot. **B**, All possible combinations of covariates were used to fit a linear model. The top 10 models, ranked by median adjusted *R*^2^ (red bars), were identified (**B**). The proportion of regressions performed by that model that were found to be statistically significant (F-test 0.05) are represented by the blue bars.

Analysis of the dependent variables included in the top performing models revealed that all models included structural protein EMP scores followed by deaths per million due to complications from COPD (90% of models). The median model size included 3 features with a maximum of 5 features and a minimum of 2 features. The model producing the best fit (median *R*^2^ = 0.791) consisted of structural protein EMP scores, gender demographics, number of deaths due to COPD complications, the proportion of the population over the age of 65, and proportion of the population that is overweight **(Figure 4B)**. All together, these results further indicate the robustness of the structural protein EMP score as a population level risk descriptor and identifies a potential candidate model for predicting pandemic severity.

## Discussion

In the present study, we uncover evidence supporting an association between population SARS-CoV-2 infection outcome and MHC-I genotype. In line with related work highlighting the relationship between total epitope load with HIV viral control^33^, we arrive at a working model that MHC-I alleles presenting more unique SARS-CoV-2 epitopes will be associated with lower mortality due to a higher number of potential T cell targets. The SARS-CoV-2 binding capacities of 52 common MHC-I alleles were assessed using the EnsembleMHC prediction platform. These predictions identified 971 high-confidence MHC-I peptides out of a candidate pool of nearly 3.5 million. In agreement with other *in silico* studies^15,34^, the assignment of the predicted peptides to their respective MHC-I alleles revealed an uneven distribution in the number of peptides attributed to each allele. We discovered that the MHC-I peptide-allele distribution originating from the full SARS-CoV-2 proteome undergoes a notable rearrangement when considering only peptides derived from viral structural proteins. The structural protein-specific peptide-allele distribution produced a distinct hierarchy of allele binding capacities. This finding has important clinical implications as a majority of SARS-CoV-2 specific CD8^+^ T cell response is directed towards SARS-CoV-2 structural proteins^8^. Therefore, patients who express MHC-I alleles enriched with a large potential repertoire of SARS-CoV-2 structural proteins peptides may benefit from a broader CD8^+^ T cell immune response.

The variations in SARS-CoV-2 peptide-allele distributions were analyzed at epidemiological scale to track its impact on country-specific mortality. Each of the 23 countries were assigned a population SARS-CoV-2 binding capacity (or EnsembleMHC population score) based on the individual binding capacities of the selected 52 MHC-I alleles weighted by their endemic population frequencies. This hierarchization revealed a strong inverse correlation between EnsembleMHC population score and observed population mortality, indicating that populations enriched with high SARS-CoV-2 binding capacity MHC-I alleles may be better protected. The correlation was shown to be stronger when calculating the EnsembleMHC population scores with respect to only structural proteins, reinforcing their relevance to viral immunity. Finally, The molecular origin of the 108 predicted peptides specific to SARS-CoV-2 structural proteins revealed that they are derived from enriched regions with a minimal predicted impact from amino acid sequence polymorphisms.

The utility of structural protein EnsembleMHC population scores was further supported by a multivariate analysis of additional SARS-CoV-2 risk factors. These results emphasized the relative robustness of structural protein EMP scores as a population risk assessment tool. Furthermore, a linear model based on the combination of structural protein EMP scores and select population-level risk factors was identified a potential candidate for a predictive model for pandemic severity. As such, the incorporation of the structural protein EMP score in more sophisticated models will likely improve epidemiological modeling of pandemic severity.

In order to achieve the highest level of accuracy in MHC-I predictions, the most up-to-date versions of each component algorithm were used. However, this meant that several of the algorithms (MHCflurry, netMHCpan-EL-4.0 and MixMHCpred) were benchmarked against subsets of mass spectrometry data that were used in the original training of these MHC-I prediction models. While this could result in an unfair weight applied to these algorithms in *peptide*^*FDR*^ calculation, the individual *FDR*s of MHCflurry, netMHCpan-EL-4.0 and MixMHCpred were comparable to algorithms without this advantage **(SI A.2)**. Furthermore, the peptide selection of SARS-CoV-2 peptides was shown to be highly cooperative within EnsembleMHC **(SI A.7)**, and individual algorithms failed to replicate the strong observed correlations between population binding capacity and observed SARS-CoV-2 mortality **(SI A.20)**.

In the future, the presented model could be applied to predict individual T cell capacity to mount a robust SARS-CoV-2 immune response. Evolutionary divergence of patient MHC-I genotypes have shown to be predictive of response to immune checkpoint therapy in cancer and HIV^35,36^. However, confirmation will require large data sets associating individual patient MHC-I genotype and outcome. Additionally, future use of EnsembleMHC to design personalized T cell vaccines will require broad experimental validation of high scoring peptides, since EnsembleMHC predicts MHC-I peptides with a high probability of antigen presentation as opposed to directly predicting peptide immunogenicity. While previous work has determined that a majority of successfully presented viral MHC-I peptides are immunogenic^37^, there is an expectation that some presented SARS-CoV-2 MHC-I peptides will fail to produce an immune response.

The current work assessed the relative importance of the structural protein EMP score with respect to other population-level risk factors (e.g. population incidence of risk-associated commodities, healthcare infrastructure, age, sex), however, it should be noted that the impacts these risk factors on patient outcome are likely to vary significantly on a individual basis. Furthermore, other genetic determinants of severity were not considered^38^. Therefore, a complete understanding of the relative importance of MHC genotype and SARS-CoV-2 presentation capacity on patient outcome will require the integration individual patient genetic and clinical data.

The versatility of the proposed model will be improved by the consideration of additional MHC-I alleles. To reduce the presence of confounding factors, EnsembleMHC was parameterized on only a subset of common MHC-I alleles that had strong existing experimental validation. While the selected MHC-I alleles are among some of the most common, personalized risk assessment will require consideration of the full patient MHC-I genotype. The continued mass spectrometry-based characterization of MHC-I peptide binding motifs will help in this regard. However, due to the large potential sequence space of the MHC-I protein, extension of this model will likely require inference of binding motifs based on MHC variant clustering.

## Data Availability

HLA frequency data was obtained from the Allele Frequency Net database.
Coronavirus pandemic information was obtained from the Center for Systems Science and Engineering (CSSE) at Johns Hopkins University.
Mass spectrometry data was obtained from the UCSD MASSive database (MSV000084442).
SARS-CoV-2 sequences were obtained from NCBI Virus database.
Country health data was obtained from the Global Health Observatory data repository provided by the World Health Organization

http://www.allelefrequencies.net

https://github.com/CSSEGISandData/COVID-19.git

https://massive.ucsd.edu/ProteoSAFe/static/massive.jsp

https://www.ncbi.nlm.nih.gov/labs/virus/vssi/#/

https://apps.who.int/gho/data/node.main

## Acknowledgments

We would like to thank Drs. Diego Chowell, Matthew Scotch, Sri Krishna, Shay Ferdosi, and Mr. John Vant, Mr. Ryan Boyd, and Ms. Mollie Peters for critical feedback and discussion. Finally, we would like to thank ASU Research computing for allocating the computational resources.

## Methods

### EnsembleMHC prediction workflow

#### EnsembleMHC component binding and processing prediction algorithms

EnsembleMHC incorporates MHC-I binding and processing predictions from 7 publicly available algorithms: MHCflurry-affinity-1.6.0^19^, MHCflurry-presentation-1.6.0^19^, netMHC-4.0^21^, netMHCpan-4.0-EL^20^, netMHCstabpan-1.0^24^, PickPocket-1.1^23^ and, MixMHCpred-2.0.2^22^. These algorithms were chosen based on the criteria of providing a free academic license, bash command line integration, and demonstrated accuracy for predicting SARS-CoV-2 MHC-I peptides with experimentally validated binding stability^39^.

Each of the selected algorithms cover components of MHC-I binding and antigen processing that roughly fall into two categories: ones based primarily on MHC-I binding affinity predictions and others that model antigen presentation. To this end, MHCflurry-affinity, netMHC, PickPocket, and netMHCstabpan predict binding affinity based on quantitative peptide binding affinity measurements. netMHCstabpan also incorporates peptide-MHC stability measurements and Pick-Pocket performs prediction based on binding pocket structural extrapolation. To model the effects of antigen presentation, MixMHCpred, netMHCpan-EL, and MHCflurry-presentation are trained on naturally eluted MHC-I ligands. Additionally, MHCflurry-presentation incorporates an antigen processing term.

#### Parameterization of EnsembleMHC using mass spectrometry data

EnsembleMHC is able to achieve high levels of precision in peptide selection through the use of allele and algorithm-specific binding affinity thresholds. These binding affinity thresholds were identified through the parameterization of each algorithm on high-quality mass spectrometry data sets^17^. The mass spectrometry data sets used for algorithm parameterization were collected in the largest single laboratory MS-based characterization of MHC-I peptides presented by single MHC allele cell lines. These characteristics significantly reduces the number of artifacts introduced by differences in peptide isolation methods, mass spectrometry acquisition, and convolution of peptides in multiallelic cell lines. An overview of the EnsembleMHC parameterization is provided in supplemental figures **(SI A.1)**.

Fifty-two common MHC-I alleles were selected for parameterization based on the criteria that they were characterized in *Sarkizova et al*. data sets and that all 7 component algorithms could perform peptide binding affinity predictions for that allele. Each target peptide (observed in the MS data set) was paired with 100 length-matched randomly sampled decoy peptides (not observed in the MS data set) derived from the same source proteins. If a protein was less than 100 amino acids in length, then every potential peptide from that protein was extracted.

Each of the seven algorithms were independently applied to each of the 52 allele data sets. For each allele data set, the minimum score threshold was determined for each algorithm that recovered 50% of the allele repertoire size (the total number of target peptides observed in the MS data set for that allele). Additionally, the expected accuracy of each algorithm was assessed by calculating the observed false detection rate (the fraction of identified peptides that were decoy peptides) using the identified algorithm and allele specific scoring threshold. The parameterization process was repeated 1000 times for each allele through bootstrap sampling of half of the peptides in each single MHC allele data set. The final *FDR* and score threshold for each algorithm at each allele was determined by taking the median value of both quantities reported during bootstrap sampling.

#### Peptide confidence assessment

Peptide confidence is assigned by calculating the *peptide*^*F D R*^. This quantity is defined as the product of the empirical *FDR*s of each individual algorithm that detected a given peptide. The *peptide*^*F D R*^ is calculated using equation 1,

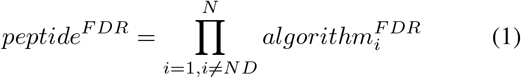

where *N* is the number of MHC-I binding and processing algorithms, *ND* represents an algorithm that did not detect a given peptide, and *algorithm*^*FDR*^ represents the allele specific *FDR* of the Nth algorithm.

The *peptide*^*FDR*^ represents the joint probability that all MHC-I binding and processing algorithms that detected a particular peptide did so in error, and therefore returns a probability of false detection. Unless otherwise stated, EnsembleMHC selected peptides based on the criterion of a *peptide*^*FDR*^ ≤ 5%.

### Application of EnsembleMHC to tumor cell line data

#### Tumor MHC-I peptide data sets

Ten tumor samples were obtained from the *Sarkizova et al*. data sets. Tumor samples were selected for analyis if at least 50% of the expressed MHC-I alleles for that sample were included in the 52 MHC-I alleles supported by EnsembleMHC. For each data set, decoy peptides were generated in a manner identical to the method used for algorithm parameterization on single MHC allele data.

#### Tumor MHC-I peptide identification

Peptide identification by each algorithm was based on restrictive or permissive binding affinities thresholds. These thresholds correspond to commonly used score cutoffs for the identification of strong binders (restrictive) or all binders (permissive). These thresholds are 0.5% (percentile rank) or 50nM (IC50 value) for strong binders, and 2% (percentile rank) or 500nM (IC50 value) for all binders. Due to the lack of recommend score thresholds for MHCflurry-presentation, the raw presentation score was converted to a percentile score by histogramming the presentation scores produced by 100,000 randomly generated peptides.

### Application of EnsembleMHC for the prediction of SARS-CoV-2 MHC-I peptides

#### SARS-CoV-2 reference sequence

MHC-I peptide predictions for the SARS-CoV-2 proteome were performed using the Wuhan-Hu-1(MN908947.3) reference sequence (https://www.ncbi.nlm.nih.gov/genbank/sars-cov-2-seqs/). All potential 8-14mer peptides (n= 67,207) were derived from the open reading frames in the reported proteome, and each peptide was evaluated by the EnsembleMHC workflow.

#### SARS-CoV-2 polymorphism analysis and protein structure visualizations

Polymorphism analysis of SARS-CoV-2 structural proteins were performed using 102,148 full length protein sequences obtained from the COVIDep database^40^. Solved structures for the E (5X29) and S (6VXX) proteins (http://www.rcsb.org/)^41^ and predicted structures for the M and N proteins^42^ were visualized using VMD^43^.

### Application of EnsembleMHC to determine population SARS-CoV-2 binding capacity

The peptides identified by the EnsembleMHC workflow were used to assess the SARS-CoV-2 population binding capacity by weighing individual MHC allele SARS-CoV-2 binding capacities by regional expression (for a schematic representation see SI A.6).

#### Population-wide MHC-I frequency estimates by country

The selection of countries included in the EnsembleMHC population binding capacity assessment was based on several criteria regarding the underlying MHC-I allele data for that country **(SI A.6)**. The MHC-I allele frequency data used in our model was obtained from the Allele Frequency Net Database (AFND)^18^, and frequencies were aggregated by country. However, the currently available population-based MHC-I frequency data has specific limitations and variances, which we have addressed as follows:

##### Quality of MHC data within countries

We define MHC-typing breadth as the diversity of identified MHC-I alleles within a given country, and its depth as the ability to accurately achieve 4-digit MHC-I genotype resolution. High variability was observed in both the MHC-I genotyping breadth and depth **(SI A.6 inset)**. Consequently, additional filter-measures were introduced to capture potential sources of variance within the analyzed cohort of countries. The thresholds for filtering the country-wide MHC-I allele data were set based on meeting two inclusion criteria: 1) MHC genotyping of at least 1000 individuals have been performed in that population, avoiding skewing of allele frequencies due to small sample size. 2) MHC-I allele frequency data for at least 51 of the 52 (95%) MHC-I alleles for which the EnsembleMHC was parameterized to predict, ensuring full power of the EnsembleMHC workflow.

##### Ethnic communities within countries

In instances where the MHC-I allele frequencies would pertain to more than one community, the reported frequencies were counted towards both contributing groups. For example, the MHC-I frequency data pertaining to the Chinese minority in Germany would be factored into the population MHC-I frequencies for both China and Germany. In doing so, this treatment resolves both ancestral and demographic MHC-I allele frequencies.

#### Normalization of MHC allele frequency data

The focus of this work was to uncover potential differences in SARS-CoV-2 MHC-I peptide presentation dynamics induced by the 52 selected alleles within a population. Accordingly, the MHC-I allele frequency data was carefully processed in order to maintain important differences in the expression of selected alleles, while minimizing the effect of confounding variables.

The MHC-I allele frequency data for a given population was first filtered to the 52 selected alleles. These allele frequencies were then converted to the theoretical total number of copies of that allele within the population (*allele count*) following

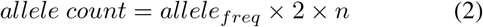

where *allele*_*freq*_ is the observed allele frequency in a population and *n* is the population sample size for which that allele frequency was measured. The allele count is then normalized with respect to the total allele count of selected 52 alleles within that population using the following relationship

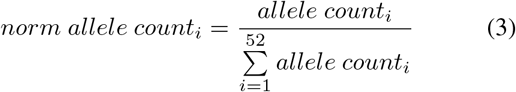

where *i* is one of the 52 selected alleles. This normalization is required to overcome the potential bias towards *hidden alleles* (alleles that are either not well characterized or not supported by EnsembleMHC) as would be seen using alternative allele frequency accounting techniques (e.g. sample-weighted mean of selected allele frequencies or normalization with respect to all observed alleles within a population **(SI A.22)**). The SARS-CoV-2 binding capacity of these *hidden alleles* cannot be accurately determined using the EnsembleMHC workflow, and therefore important potential relationships would be obscured.

#### EnsembleMHC population score

The predicted ability of a given population to present SARS-CoV-2 derived peptides was assessed by calculating the EnsembleMHC Population (EMP) score. After the MHC-I allele frequency data filtering steps, 23 countries were included in the analysis. The calculation of the EnsembleMHC population score is as follows

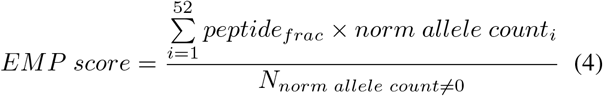

where *norm allele count* is the observed normalized allele count for a given allele in a population, *N*_*norm allele count* =0_ is the number of the 52 select alleles detected in a given population (range 51-52 alleles), and *peptide*_*frac*_ is the peptide fraction or the fraction of total predicted peptides expected to be presented by that allele within the total set of predicted peptides with a *peptide*^*FDR*^ ≤ 5%.

#### Death rate-presentation correlation

The correlation between the EMP score and the observed deaths per million within the cohort of selected countries was calculated as a function of time. SARS-Cov-2 data covering the time dependent global evolution of the SARS-CoV-2 pandemic was obtained from Johns Hopkins University Center for Systems Science and Engineering^44^ covering the time frame of January 22nd to April 9th. The temporal variations in occurrence of community spread observed in different countries were accounted for by rescaling the time series data relative to when a certain minimum death threshold was met in a country. This analysis was performed for minimum death thresholds of 1-100 total deaths by day 0, and correlations were calculated at each day sequentially following day 0 until there were fewer than 8 countries remaining at that time point. The upper-limit of 100-deaths was chosen to ensure availability of death-rate data on at least 50% of the countries for a minimum of 7 days starting following day 0. Additionally, a steep decline in average statistical power is observed with day 1 death thresholds greater than 100 deaths **(SI A.23)**.

The time death correlation was computed using Spearman’s rank correlation coefficient (two-sided). This method was chosen due to the small sample size and non-normality of the underlying data **(SI A.24)**. The reported correlations of EMP score and deaths per million using other correlation methods can be seen in supplemental figure **SI A.25**.

The low statistical power for some of the obtained correlations were addressed by calculating the Positive Predictive Value (PPV) of all correlations using the following equation^45^

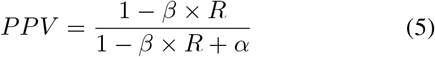

,where 1 − *β* is the statistical power of a given correlation, *R* is the pre-study odds, and *α* is the significance level. A PPV value of ≥ 95% is analogous to a p value of ≤ 0.05. Due to an unknown pre-study odd (probability that probed effect is truly non-null), R was set to 1 in the reported correlations. The proportion of reported correlations with a PPV of 95% at different R values can be seen in supplemental figure **SI A.17**. The significance of partitioning high risk and low risk countries based on median EMP score was determined using Mann-Whitney U-test. Significance values were corrected for multiple tests using the Benjamini-Hochberg procedure^30^.

#### Sub-sampling of peptides from the Full SARS-CoV-2 proteome

108 unique peptides, derived from the Full SARS-CoV-2 proteome and passing the 5% *peptide*^*FDR*^ filter, were randomly sampled. Then, the time series EMP score -death per million correlation analysis used to generate **Figure 3** was applied to each sampled peptide set. The sub-sampling procedure was repeated for 1,000 iterations **(SI A.18A)**. To quantitatively describe the similarity of the distributions, the Kullback-Leibler divergence (KLD), a measure of divergence between two probability distributions, was calculated for the correlation distribution of each sub-sample iteration relative to either the correlation distribution of the Full SARS-CoV-2 proteome or SARS-CoV-2 structural proteins **(SI A.18B)**.

### Analysis of additional SARS-CoV-2 risk factors

#### Additional SARS-CoV-2 risk factors

Twelve potential SARS-CoV-2 risk factors **(table B.4)** were selected for analysis. Country-specific data for each risk factor was obtained from the Global Health Observatory data repository provided by the World Health Organization (https://apps.who.int/gho/data/node.main). Countries were selected for analysis based on the criteria of having reported data in the WHO data sets and inclusion in the set of 23 countries for which EnsembleMHC population scores were assigned **(table B.4A)**. Data regrading the total number of noncommunicable disease-related deaths (Cardiovascular disease, Chronic obstructive pulmonary disease, and Diabetes mellitus) were converted to deaths per million.

#### Correlation of additional risk factors with observed deaths per million

Correlation analysis of each additional factor was carried out in a similar manner to that of the EnsembleMHC population score. In short, Spearman’s correlation coefficient between each individual factor and observed deaths per million was estimated as a function of time from when a specified minimum death threshold was met **(Figure 4)**. The significance level was set to *p* ≤ 0.05 and significant PPV was set to *PPV* ≥ 0.95 (eq 8).

#### Linear models of SARS-CoV-2 mortality

For the single and combination models, individual linear models were constructed for each considered death threshold as a function of time (similar to the univariate correlation analysis). Each model consisted of 1 (a single socioeconomic or health-related risk factor) or 2 (a combination of 1 risk factor and structural protein EMP score) dependent variables and deaths per million as the independent variable. The adjusted *R*^2^ value and statistical significance of the model (F-test) were then extracted from each individual model and aggregated by dependent variable **(figure 4**, **SI A.21)**.

The best performing models were determined by assessing all possible combinations of factors including structural protein EMP score. This resulted in the consideration of 4,083 different linear models. The top performing models were then selected by ranking each model by median adjusted *R*^2^.

### Code and data availability

All data analysis and statistical tests were performed using the R Statistical Computing Environment v.3.6.0 (http://www.rproject.org). Data sets and example code are available at https://github.com/eawilson-CompBio/EnsembleMHC-Covid.git

## A Supplemental figures

**Figure A.1:**
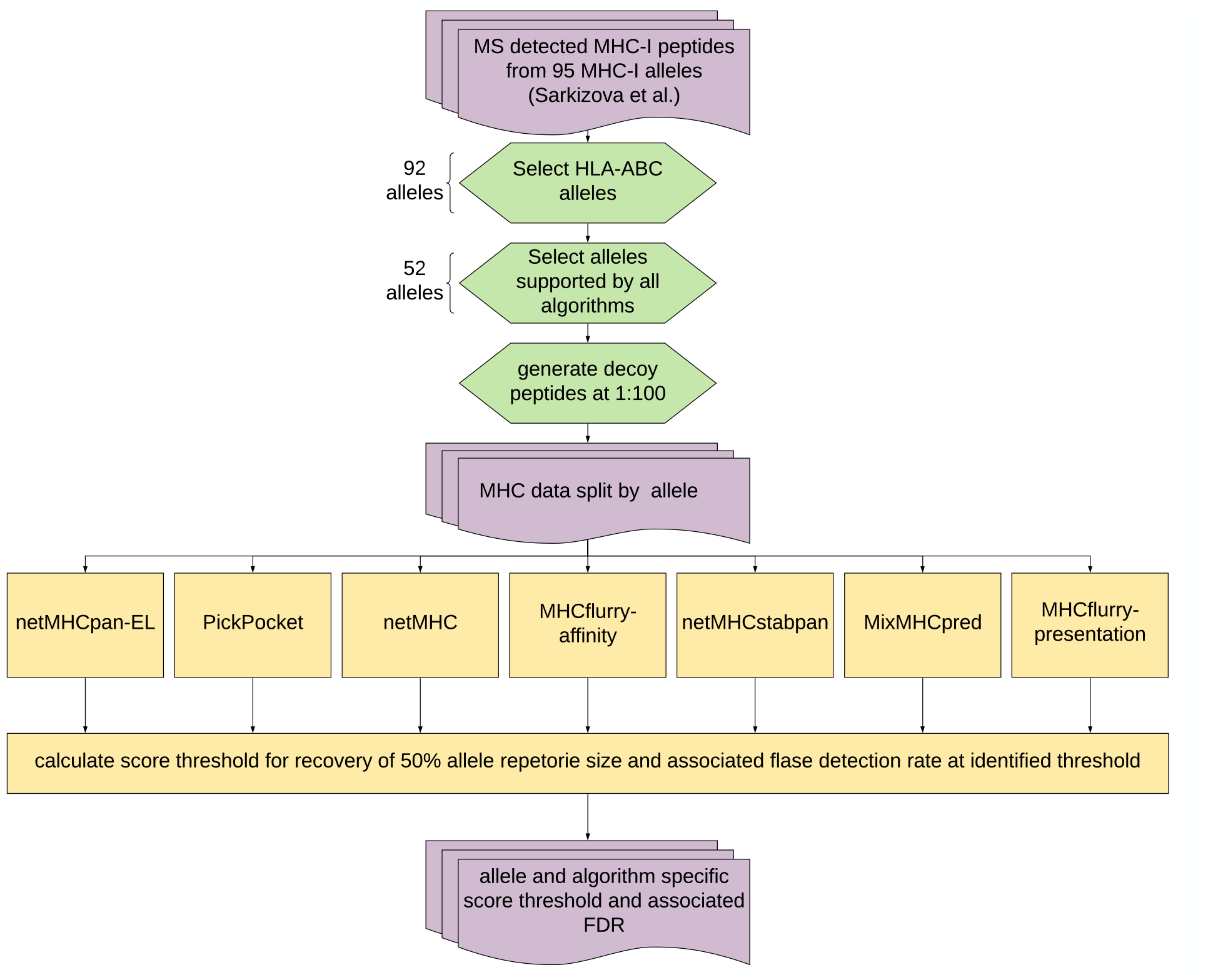
EnsembleMHC Parameterization overview.

**Figure A.2:**
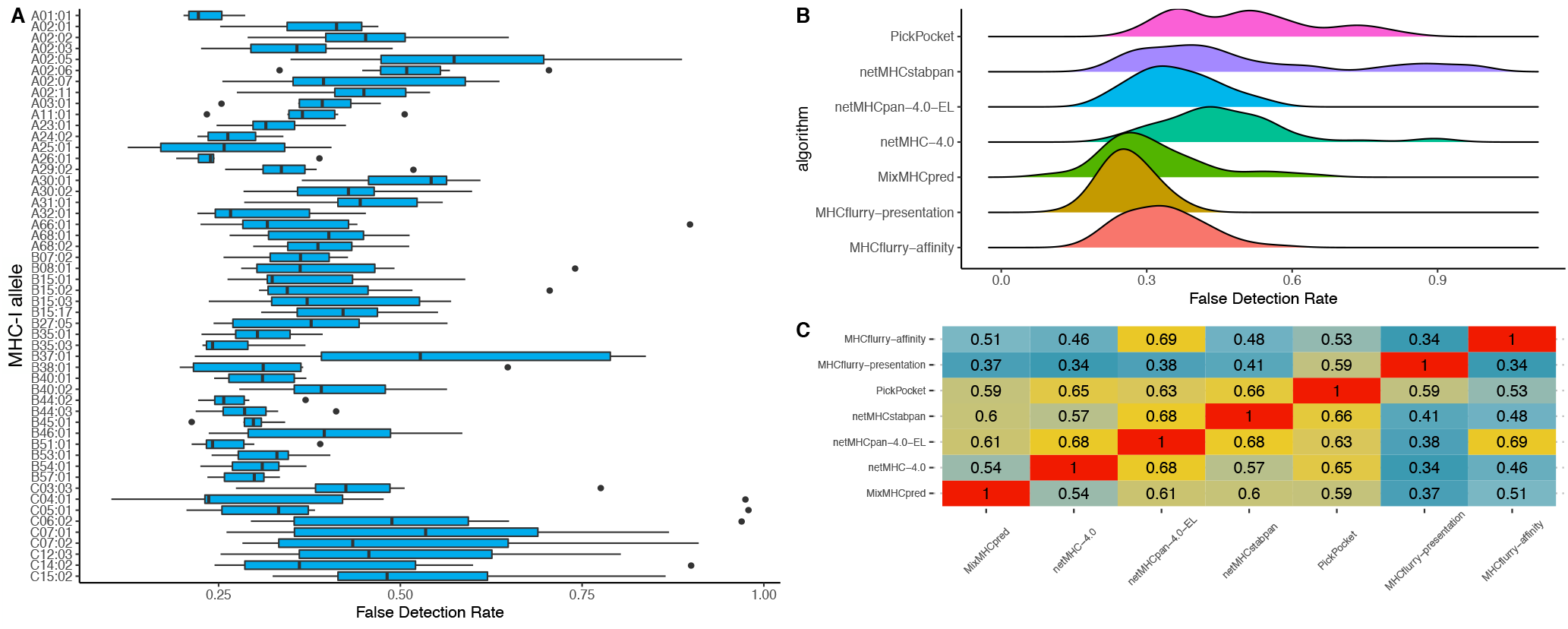
EnsembleMHC prediction workflow. **A,** The EnsembleMHC score algorithm was parameterized using high quality mass spectrometry-detected MHC-I peptides paired with a 100-fold excess of randomly generated decoy peptides. Each bar represents the distribution of algorithm-specific false detection rates (n = 7) at that MHC allele. Each box plot is in the style of Tukey. **B**, a density plot of the observed *FDR*s for each algorithm across all alleles (n = 52). **C**, The correlation between individual peptide scores for each algorithm across all alleles was calculated using Pearson correlation. Warmer colors indicate a higher level of correlation while cooler colors indicate lower correlation.

**Figure A.3:**
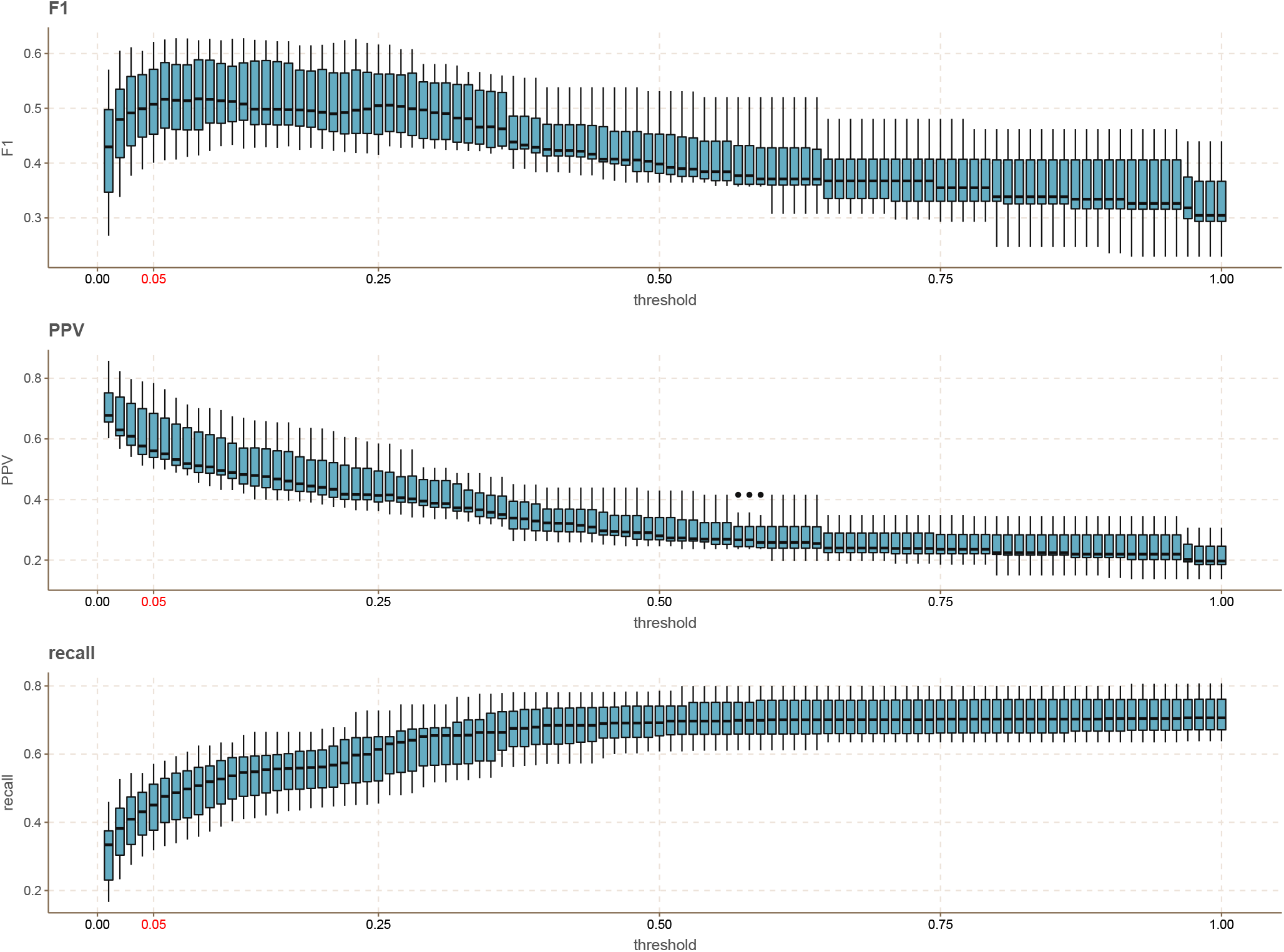
The effect of different peptide *FDR* threshold levels. The effect of different *peptide*^*FDR*^ cutoff thresholds on the results reported in **figure 1** was evaluated for a range of 0.01-1. The *peptide*^*FDR*^ selected for use in this study is highlighted in red.

**Figure A.4:**
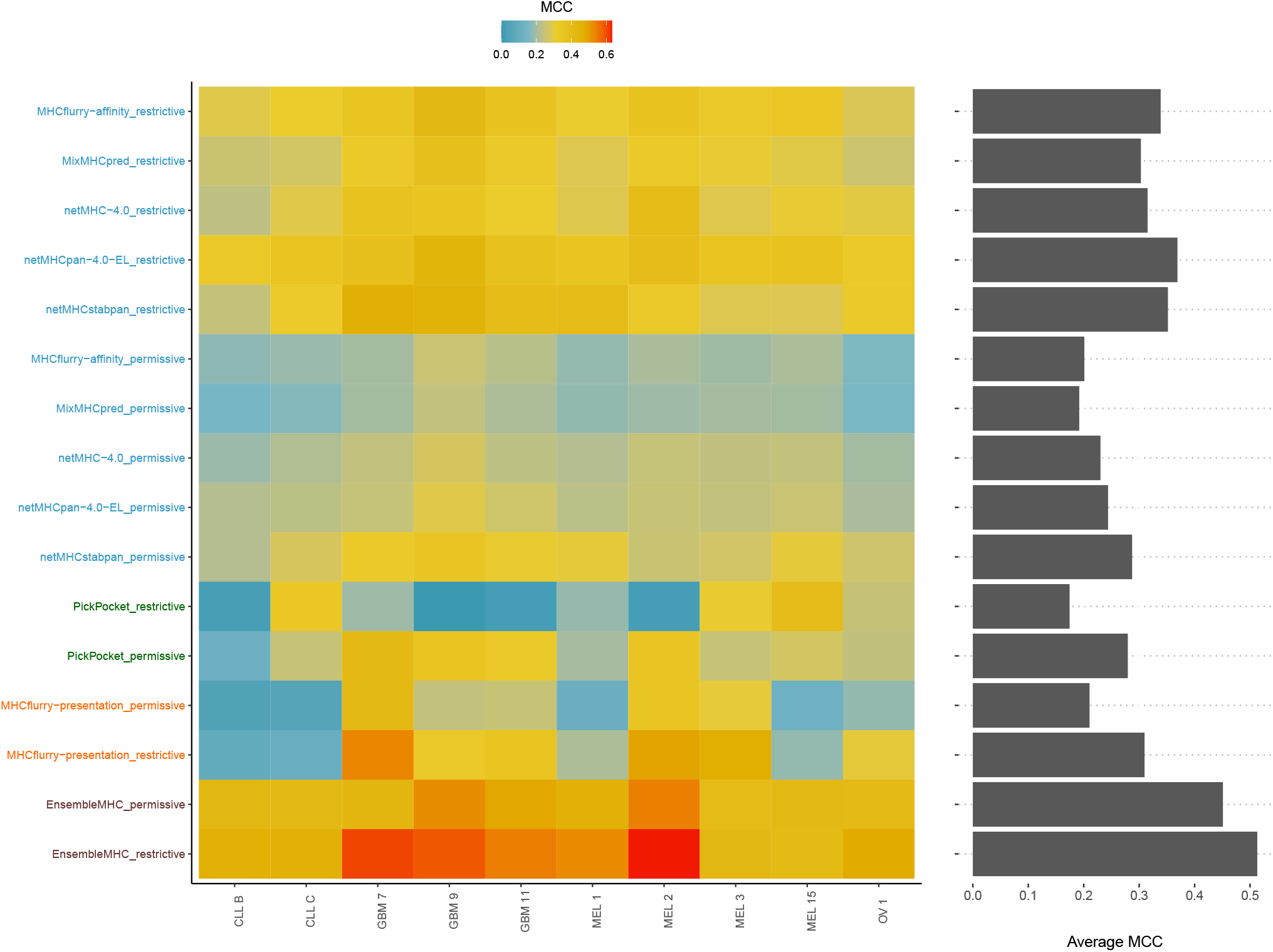
Evaluation of individual algorithms using Matthew’s correlation coefficient. As an alternative to F1 score (**Figure 1B**), Matthew’s correlation coefficient was calculated for each algorithm. Warm colors indicate higher MCC while cooler colors indicate lower MCC. The average MCC for each algorithm is represented by the margin bar plot on the right.

**Figure A.5:**
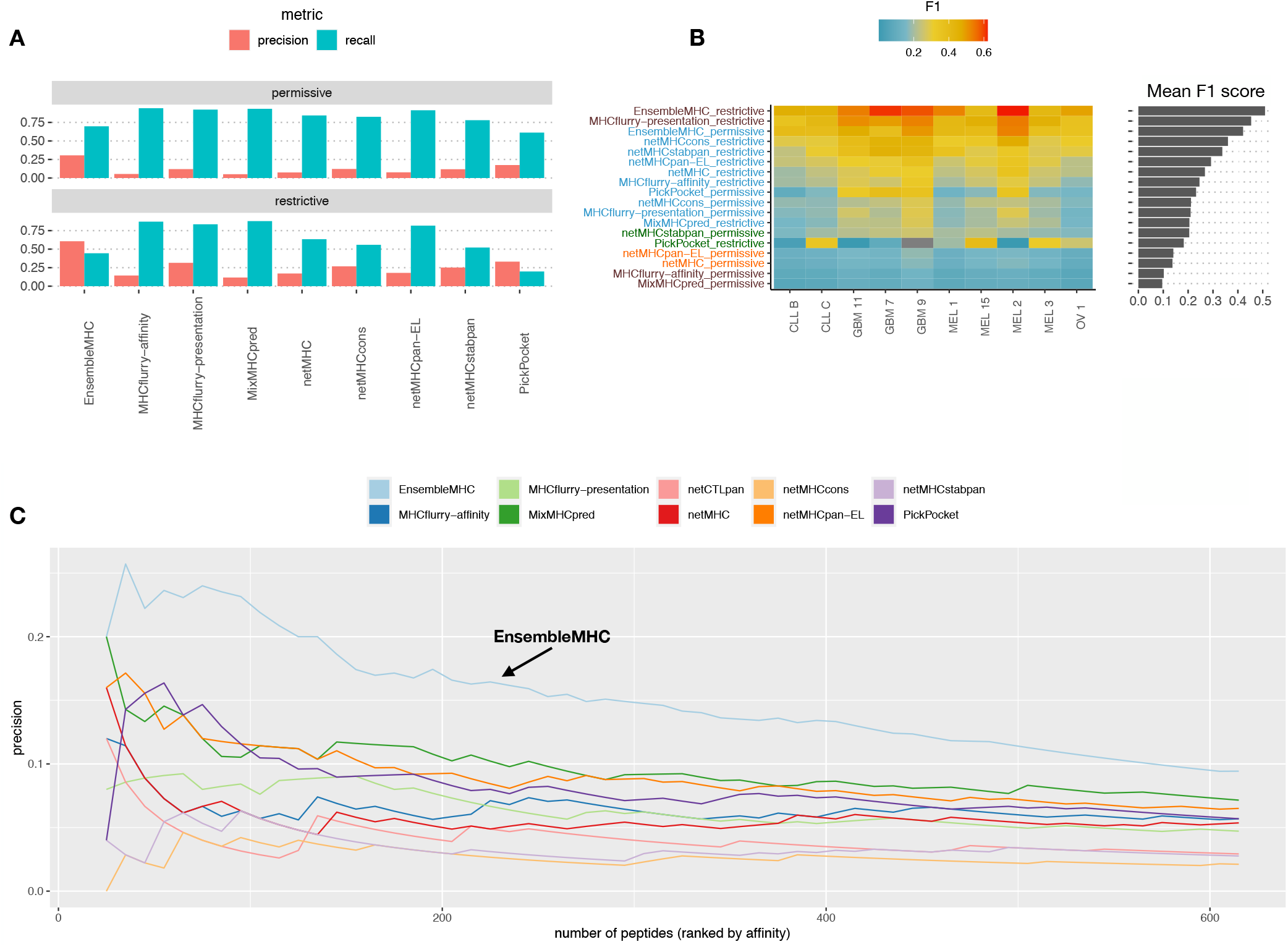
Prediction of viral immunogenic epitopes across ensemble-based algorithms. **A-B,** The results in reported **figure 1 (A-B)** were compared across ensemble-based MHC-I prediction algorithms. **C**, The positive predictive value (precision) of the algorithms to select immunogenic MHC-I peptides was assessed across Hepatitis-C genome polyprotein (P26664), Dengue virus genome polyprotein (P14340), and the HIV-1 POL-GAG protein (P03369). All potential 8 – 14mer peptides were extracted from each protein and the resulting peptides were checked against the Immune Epitope database to identify peptides with experimentally validated immunogenicity. The result of this analysis was the generation of a data set comprised of 616 experimentally validated immunogenic peptides and 54,663 putative non-immunogenic peptides. The performance of each algorithm was then assessed by calculating the precision when selecting n number of top scoring peptides as determined by a given algorithm. Precision was calculated for each algorithm across a range of *n* = 25 to *n* = 615.

**Figure A.6:**
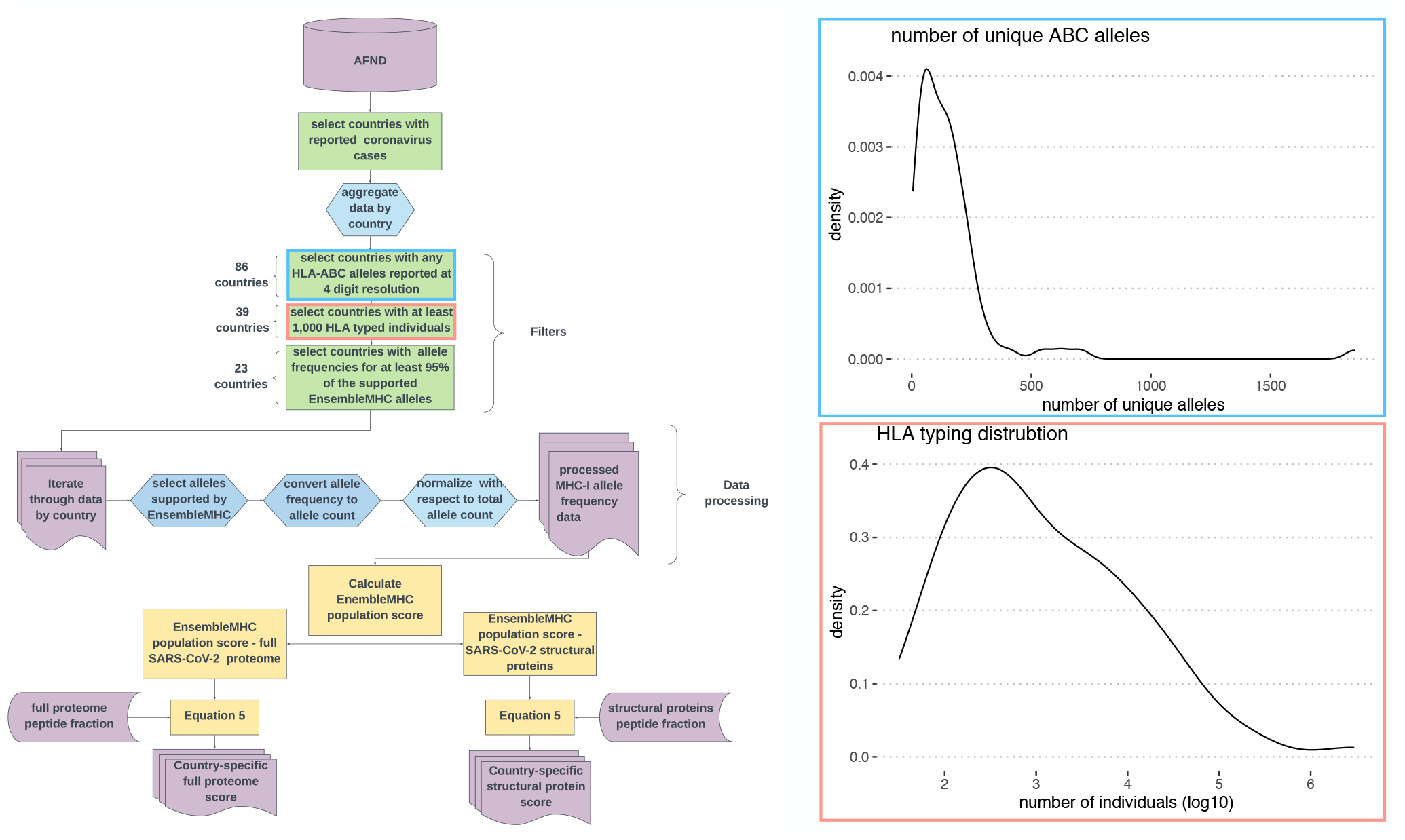
Data processing EnsembleMHC population score calculation. The overview of the data processing steps for the global MHC-I allele frequency data and its application in the calculation the EnsembleMHC population score with respect to the full SARS-CoV-2 proteome and SARS-CoV-2 structural proteins. **(inset plots)**, The blue inset plot illustrates MHC-typing breadth and depth variation by showing the distribution of the total number of MHC-I alleles reported at 4-digit resolution in 86 countries. The red inset plot shows the distribution of the number of MHC-genotyped individuals in the set of countries with at least 1 reported coronavirus case. **AFND = Allele Frequency Net Database**

**Figure A.7:**
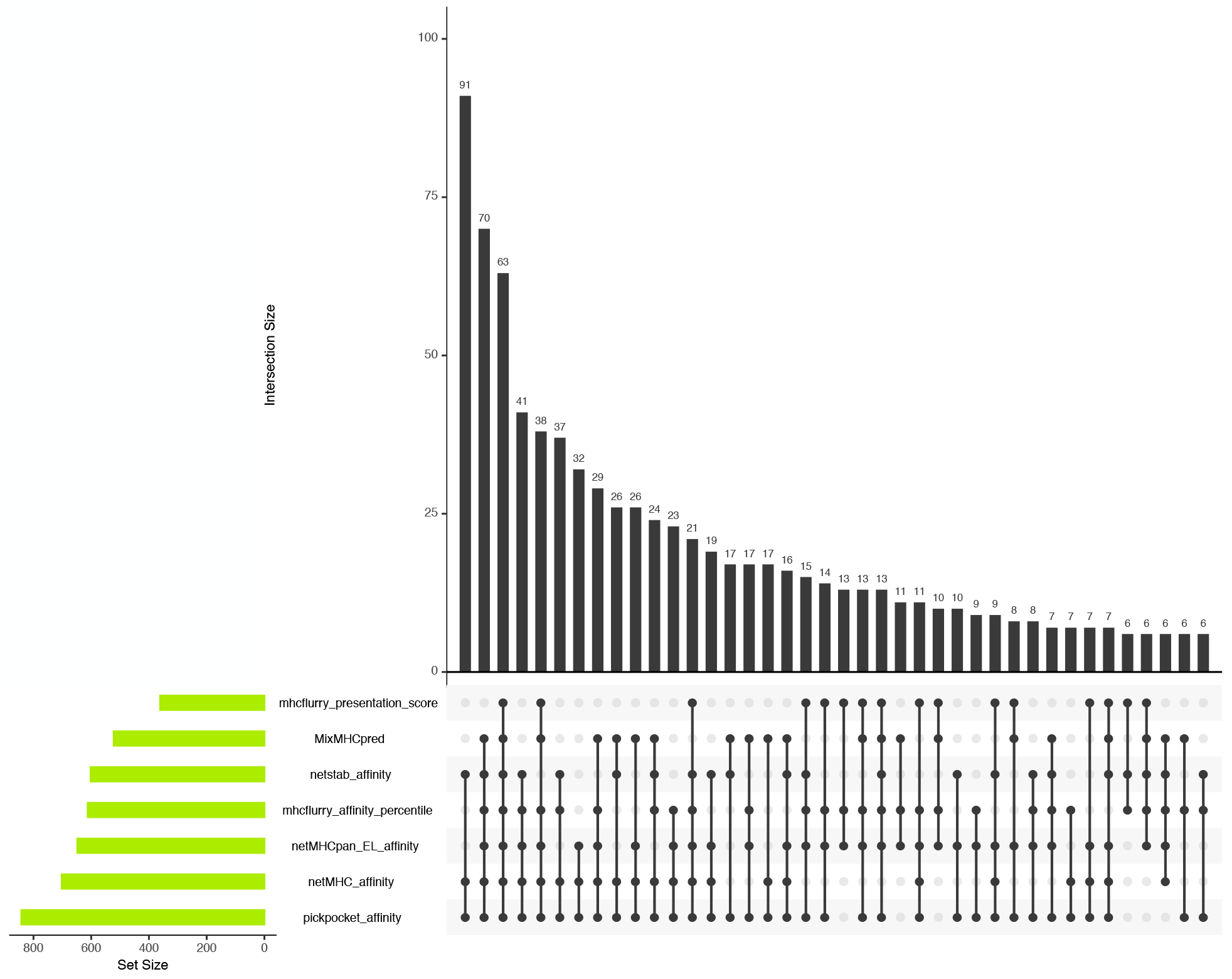
Contribution of each EnsembleMHC component algorithm to peptide selection for predicted SARS-CoV-2 peptides. The UpSet plot shows the contribution of each individual component algorithm to the 658 unique SARS-CoV-2 peptides identified by EnsembleMHC. The top bar plot indicates the number of unique peptides identified by the combination of algorithms shown by the points and segments located under each bar. The bar plot on the left-hand side of the plot indicates the total number of peptides identified by each algorithm.

**Figure A.8:**
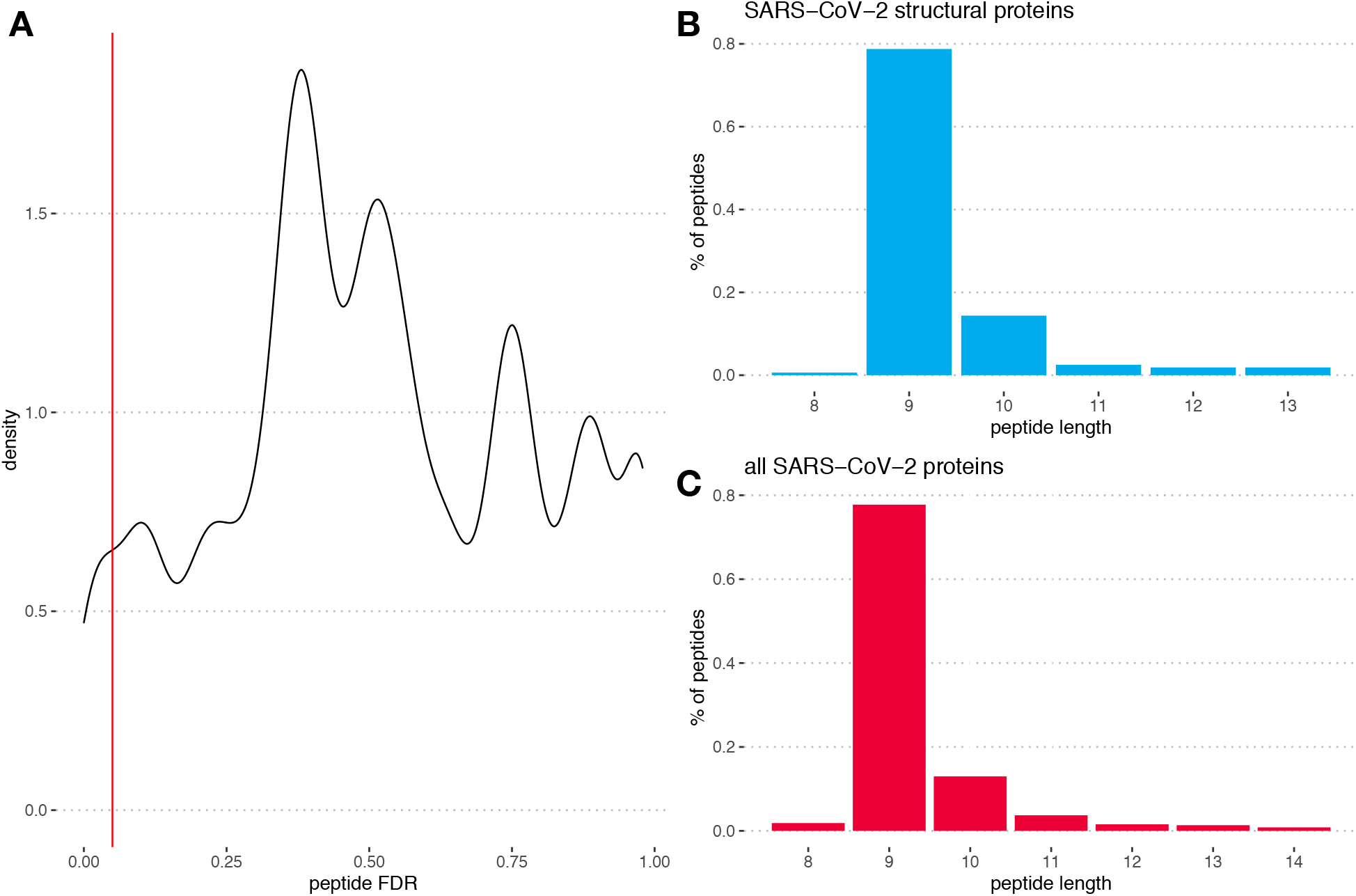
EnsembleMHC peptide^*FDR*^ and length distributions of predicted SARS-CoV-2 MHC-I peptides. **A,** The distribution of the for the *peptide*^*FDR*^ for 9,712 SARS-CoV-2 peptides that fell with the score threshold of at least one component algorithm. The red line indicates an *peptide*^*FDR*^ level of ≤ 5%. **B**, The length distribution of the 108 high-confidence peptides identified from SARS-CoV-2 structural proteins. **C**, The length distribution of the 658 high-confidence peptides identified from full SARS-CoV-2 proteome.

**Figure A.9:**
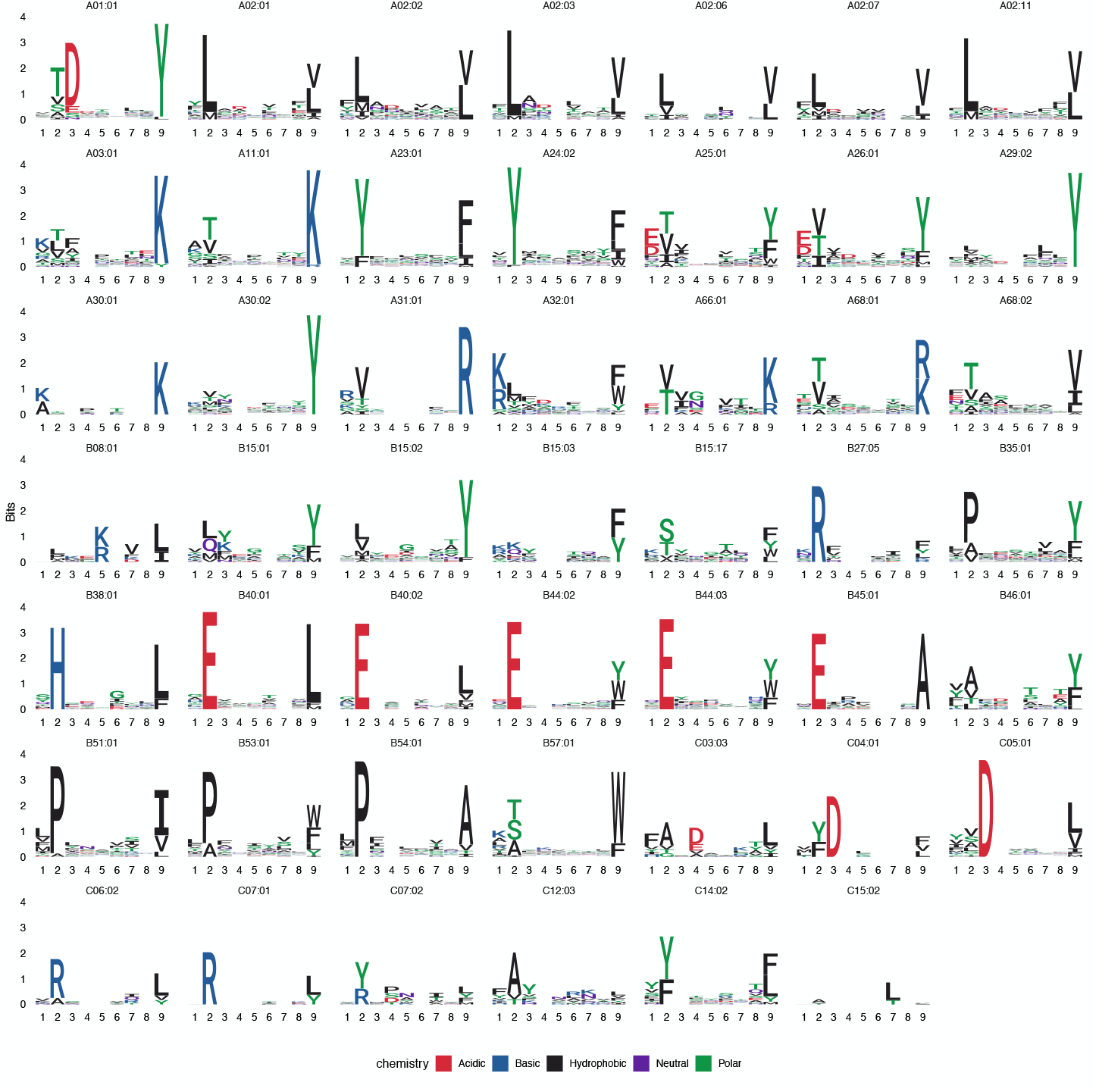
Logo plots for the identified peptides from the SARS proteome. Logo plots were generated for MHC alleles with at least 5 peptides identified by EnsembleMHC prediction. Peptides shorter than 9 amino acids had random amino acid inserted into a non-anchor position while peptides longer than 9 amino acids had a random non-anchor position deleted. Large amino acid character height indicates a high frequency of that amino acid at that position. Amino acids are colored residue type.

**Figure A.10:**
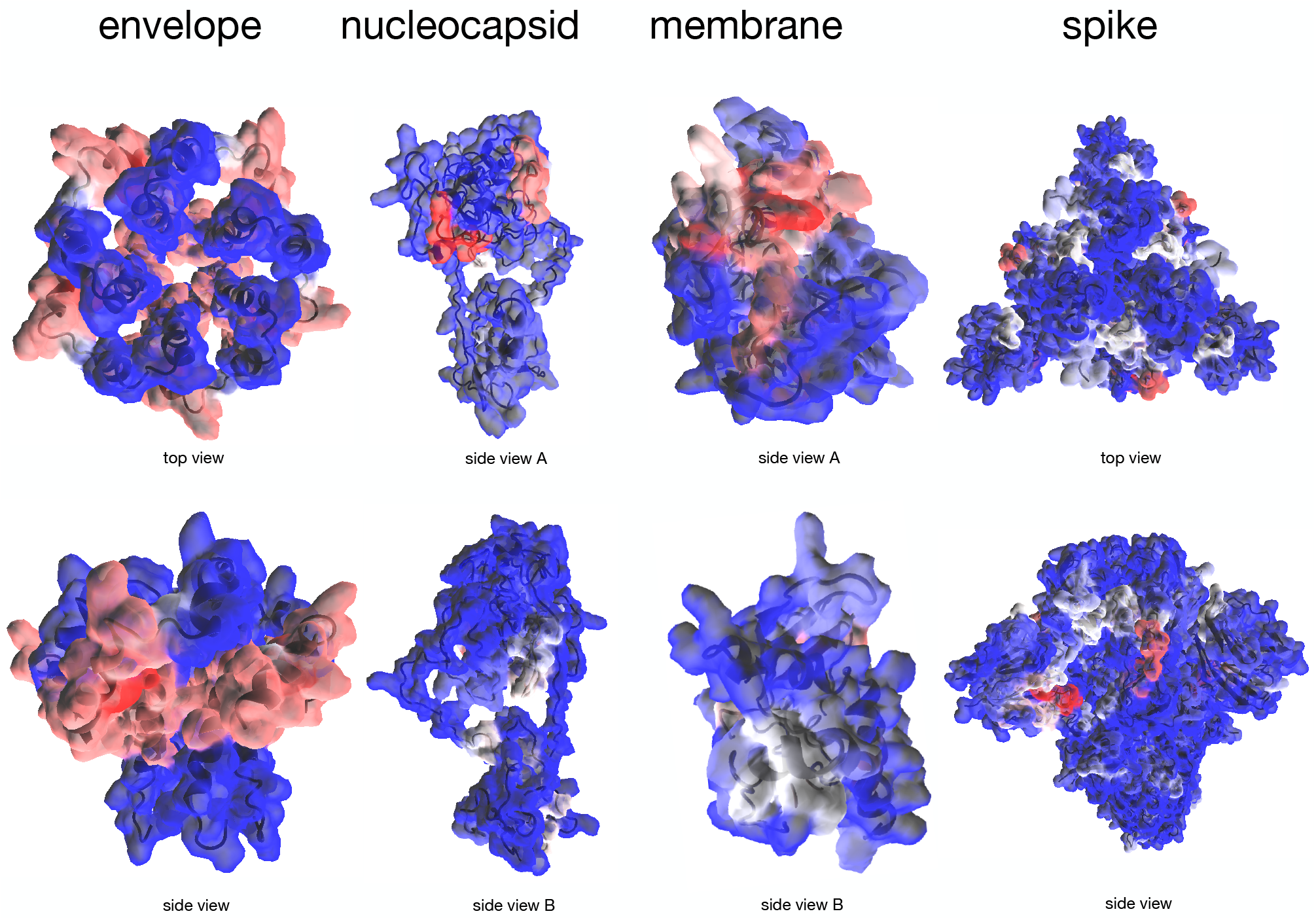
Molecular origin of predicted SARS-CoV-2 structural protein MHC-I peptides. The predicted SARS-CoV-2 structural protein MHC-I peptides were mapped onto the solved structures for the envelope and spike proteins, and the predicted structures for the nucleocapsid and membrane proteins. Red highlighted regions indicate an enrichment of predicted peptides while blue regions indicate a depletion of predicted peptides.

**Figure A.11:**
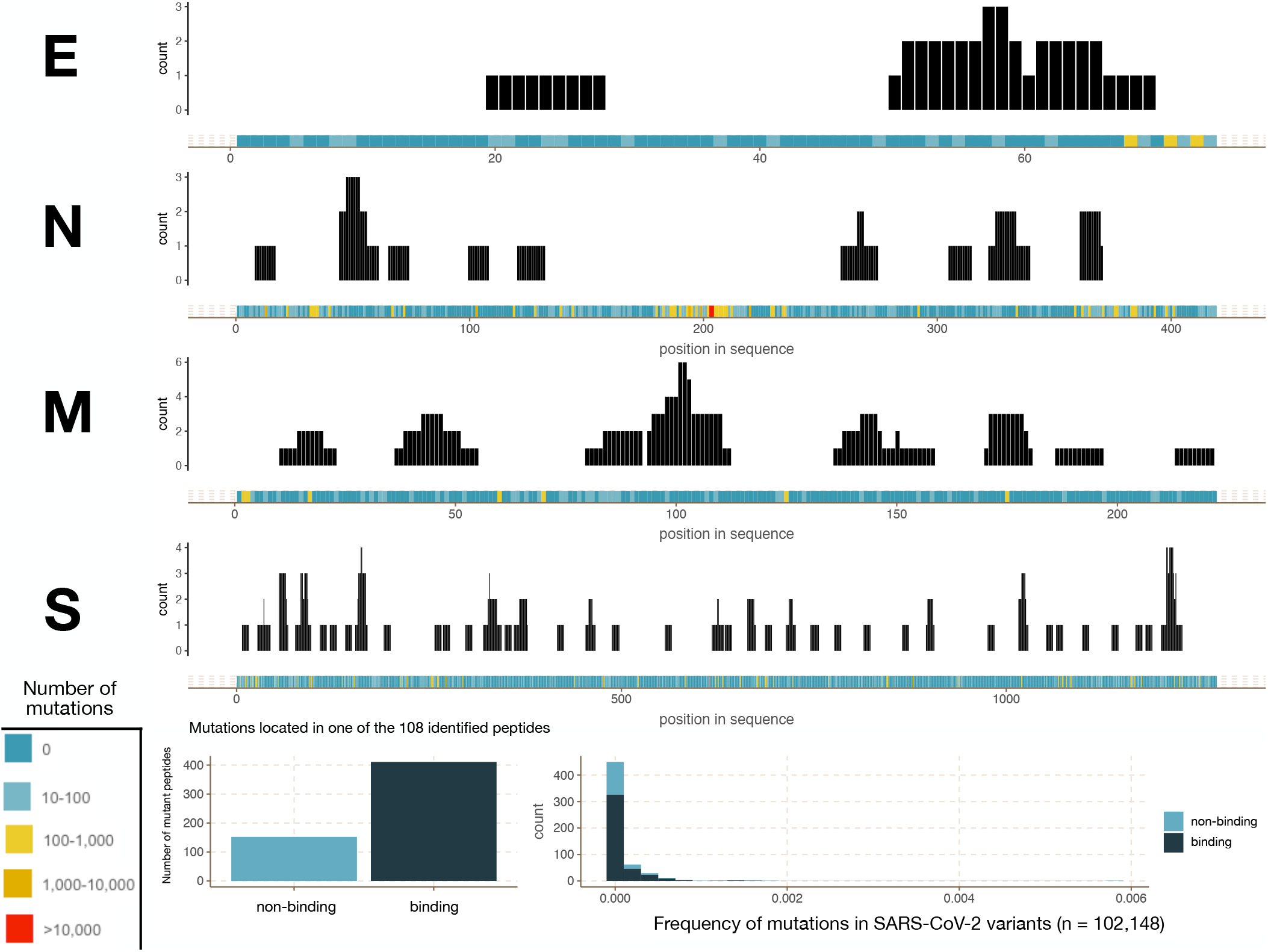
Impact of SARS-CoV-2 sequence polymorphism on predicted SARS-CoV-2 structural MHC-I peptides. Top 4 panels, The incidence of sequence position mutations (colored bar) and the frequency of each amino acid position in one of the 108 SARS-CoV-2 structural protein peptides (black bars) by protein were calculated for 102,148 SARS-CoV-2 sequence variants. **Lower left panel**, all potential mutations arising in an EnsembleMHC-predicted MHC-I peptides were evaluated for changes in binding affinity (*peptide*^*FDR*^ *>* 0.05). **Lower right panel**, The overall frequency of mutations impacting EnsmebleMHC-predicted peptides with light blue indicating deleterious mutations, and dark blue indicating neutral mutations.

**Figure A.12:**
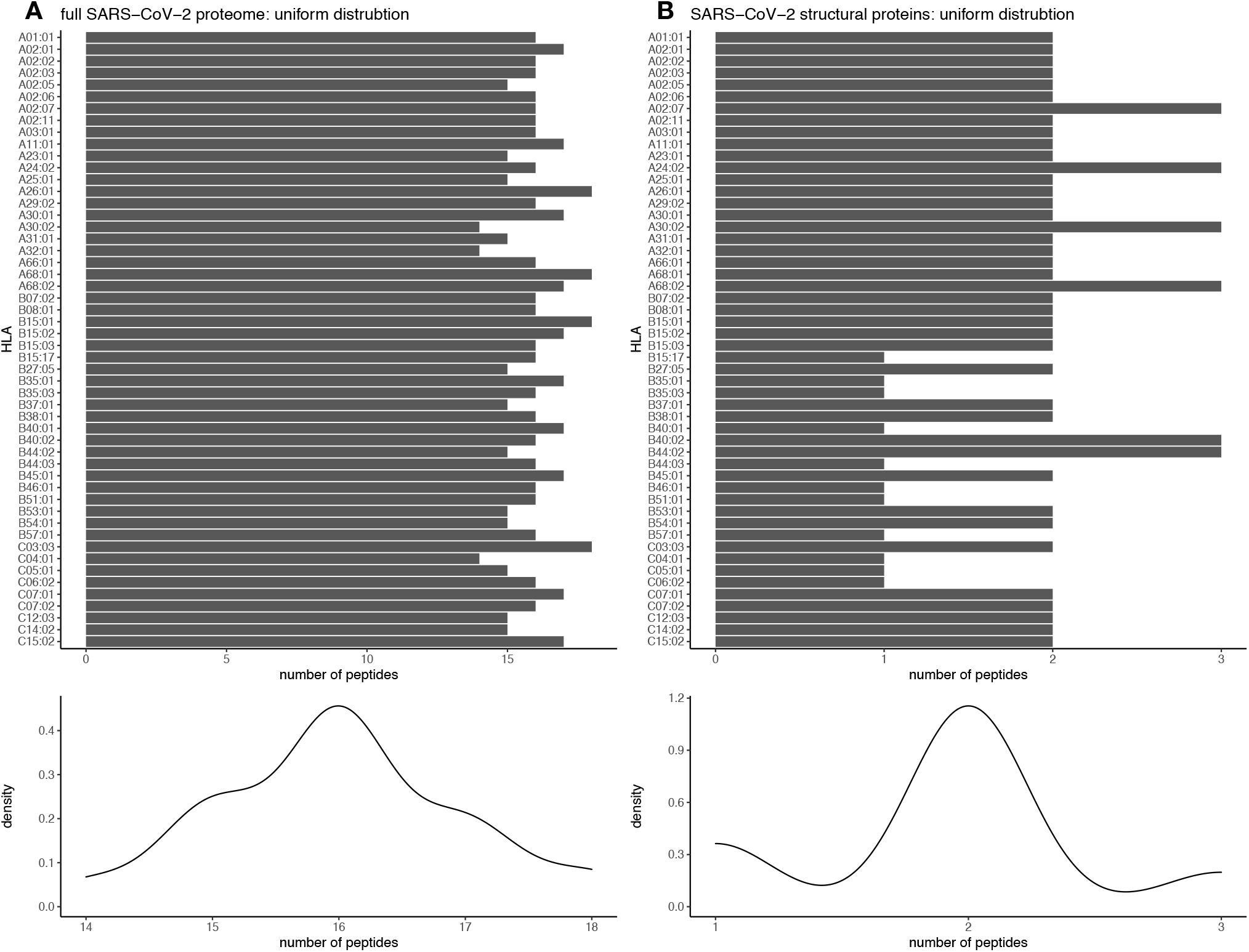
Simulated even peptide-allele distribution. The resulting SARS-CoV-2 peptide-MHC allele distrubtion was compared to a even distribution by means of the Kolmogorov-smirnov test. **A**, An even peptide-allele distribution for the full SARS-CoV-2 proteome was simulated by sampling from a discrete symmetrical distribution centered around the median number of peptides assigned to individual alleles 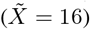. **B**, An even peptide-allele distribution for the SARS-CoV-2 structural proteins was simulated by sampling from a discrete symmetrical distribution centered around the median number of peptides assigned to individual alleles 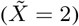.

**Figure A.13:**
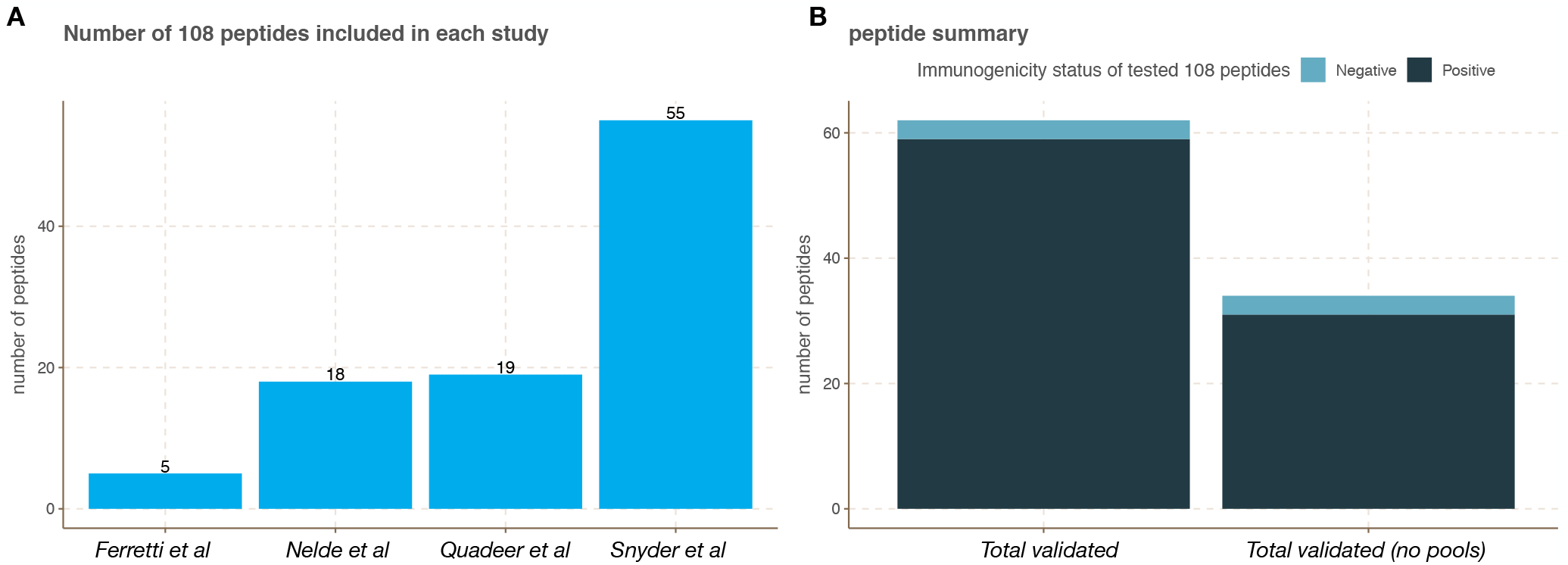
External experimental validation of the 108 high confidence SARS-Cov-2 structural protein peptides. Experimentally validated immunogenic peptides derived from SARS-CoV-2 structural proteins were obtained from 4 independent studies (Ferretti et al^46^, Nelde et al^47^, and Snyder et al^48^) and 1 meta-study (Quadeer et al^49^). These peptides were then assessed for overlap with the 108 SARS-CoV-2 peptides identified by EnsembleMHC (*108 peptides*). **A**, The total number of peptides from the *108 peptides* set that were included for testing in each study. **B**, The summary of immunogenicity status of *108 peptides* across all studies. These summaries were split into two groups. *Total validated* indicates the total number of experimentally validate 108 peptides while *total validated (no pools)* indicates the number of experimentally validated peptides excluding those only tested in peptide pools. This distinction was made due to the potential of peptide pools to obscure which tested peptide is truly responsible for the observed immune response. Overall, 57% of the predicted 108 structural protein peptides were tested with 95% of tested peptides producing an immune response.

**Figure A.14:**
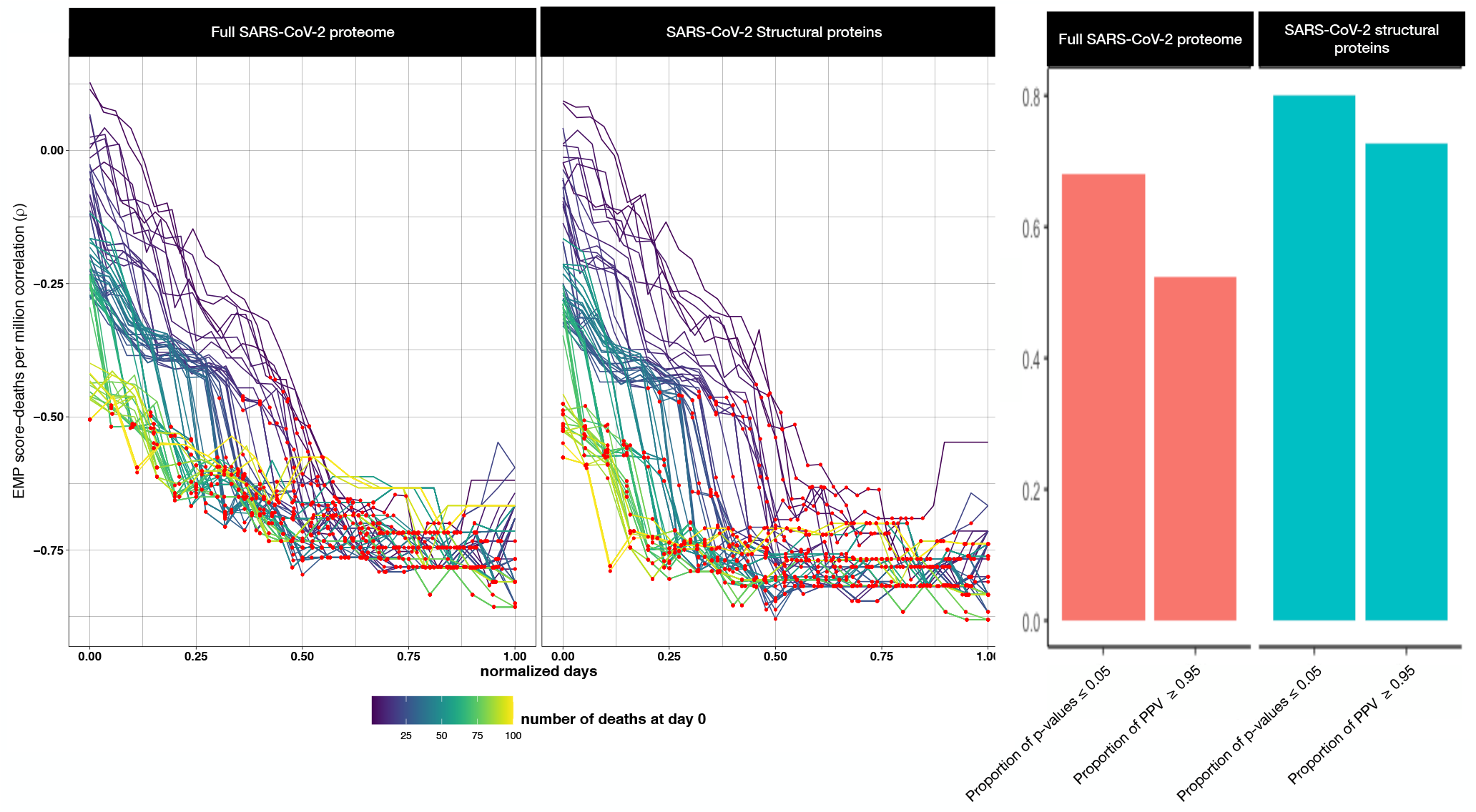
Correlation of EMP score based on full SARS-CoV-2 proteome or SARS-CoV-2 structural proteins with observed deaths per million. **A,** The correlations between EnsembleMHC population score based on the full SARS-CoV-2 proteome **(left)** or EnsembleMHC population score based on SARS-CoV-2 structural proteins **(right). B**, The difference in the proportions of significant p-values and PPV between the full SARS-CoV-2 proteome **(left)** and SARS-CoV-2 structural proteins **(right)** (not corrected for multiple testing).

**Figure A.15:**
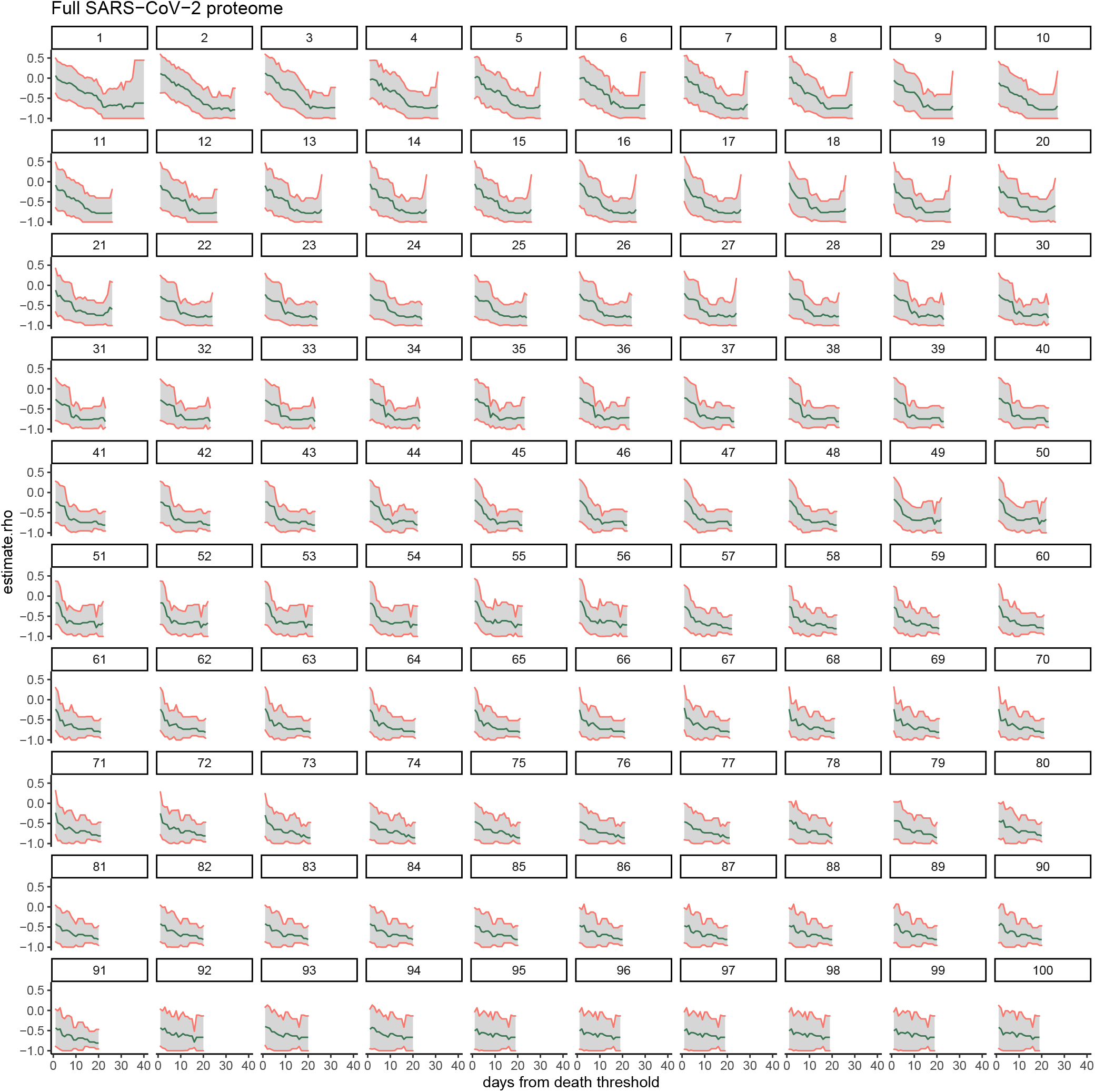
95% Confidence interval for the correlations between the EnsembleMHC score based on the full SARS-CoV-2 proteome and observed deaths per million. Each individual plot shows the 95% confidence interval (grey region) for the correlations between EMP scores based on the full SARS-CoV-2 proteome and observed deaths per million (blue line) for all starting minimum death thresholds (indicated by number above plot).

**Figure A.16:**
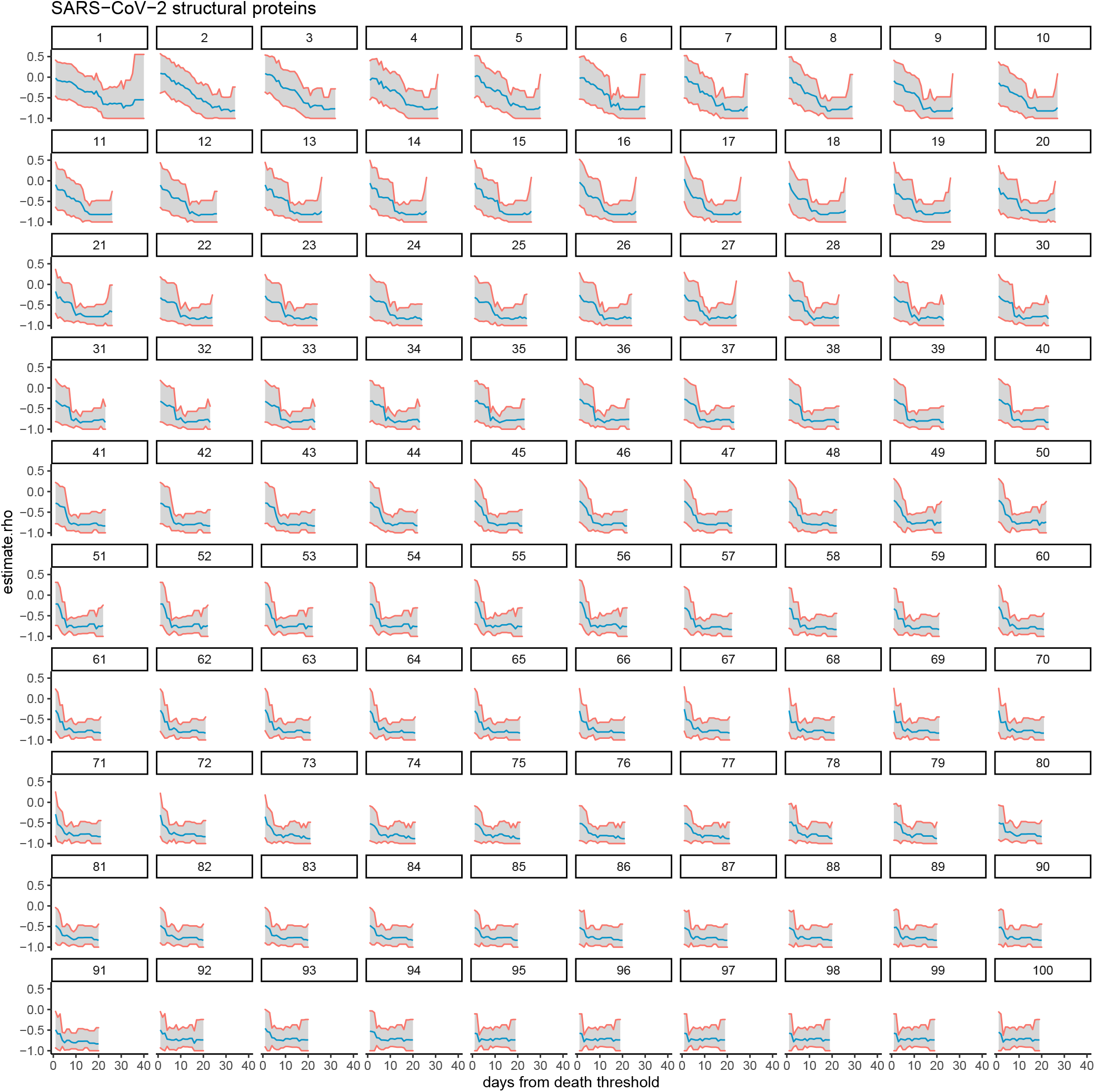
95% Confidence interval for the correlations between the EnsembleMHC score based on SARS-CoV-2 structural proteins and observed deaths per million. Each individual plot shows the 95% confidence interval (grey region) for the correlations between EMP scores based on ARS-CoV-2 structural proteins and observed deaths per million (blue line) for all starting minimum death thresholds (indicated by number above plot).

**Figure A.17:**
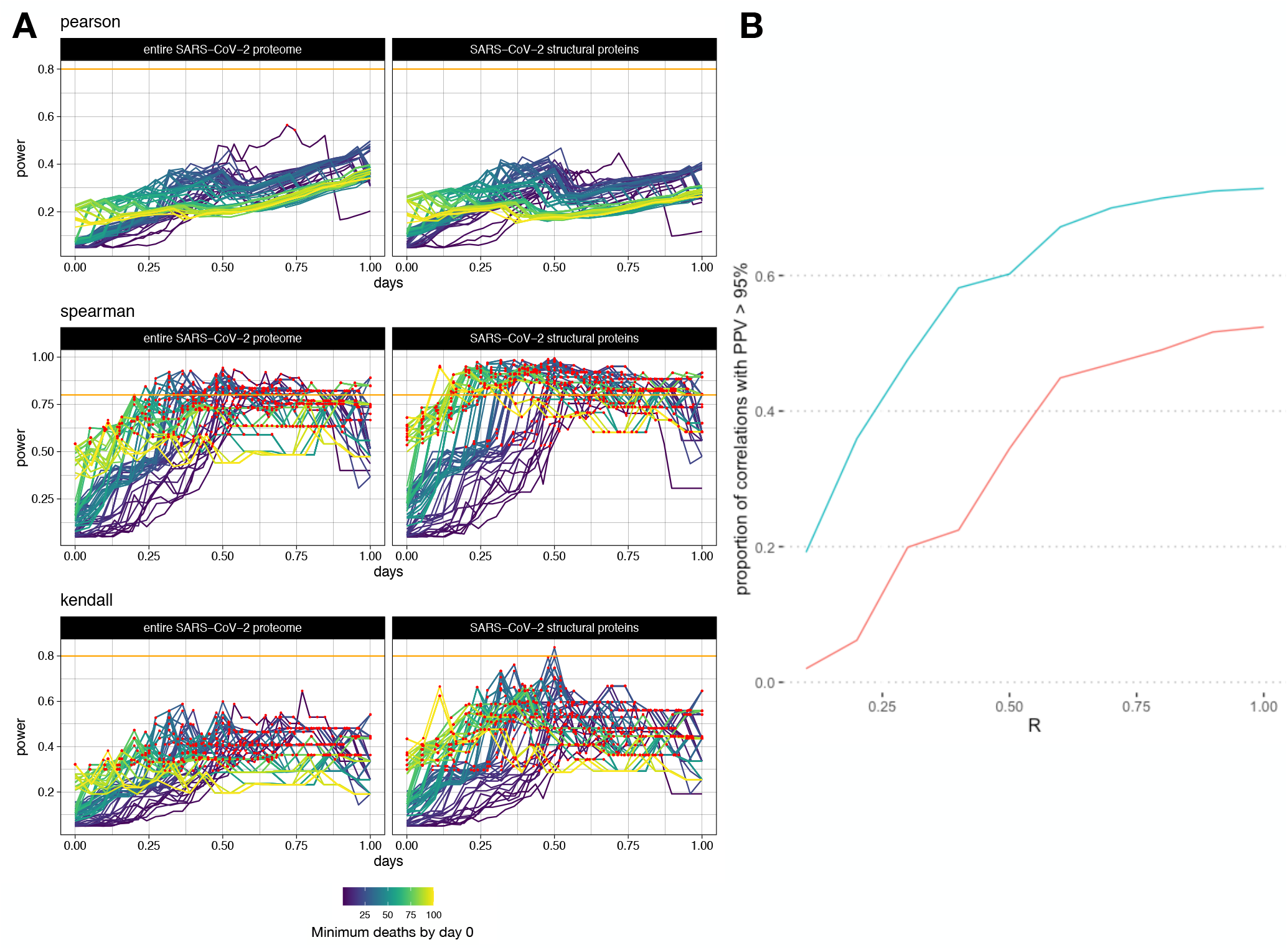
Analysis of statistical power for different correlation methods. **A,** The statistical power of each reported correlation between EnsembleMHC population score with respect to the full SARS-CoV-2 proteome (left column) or specifically structural proteins (right column) and deaths per million were calculated at each day starting from the day a country passed a particular minimum death threshold using Pearson’s correlation **(top)**, Spearman’s rho **(middle)**, and Kendall’s tau **(bottom)**. The days from each start point were normalized, and correlations that were shown to be statistically significant are colored with a red point. The orange line indicates a power threshold of 80%. **B**, The line plot on the right shows the proportion of points achieving a significant PPV at different thresholds for pre-study odds (R) for the spearman correlation carried out in section 3. The blue line represents the proportion of significant correlations for EnsembleMHC score based on SARS-CoV-2 structural proteins while the red line represents the same correlations with an EnsembleMHC population score based on the full SARS-CoV-2 proteome.

**Figure A.18:**
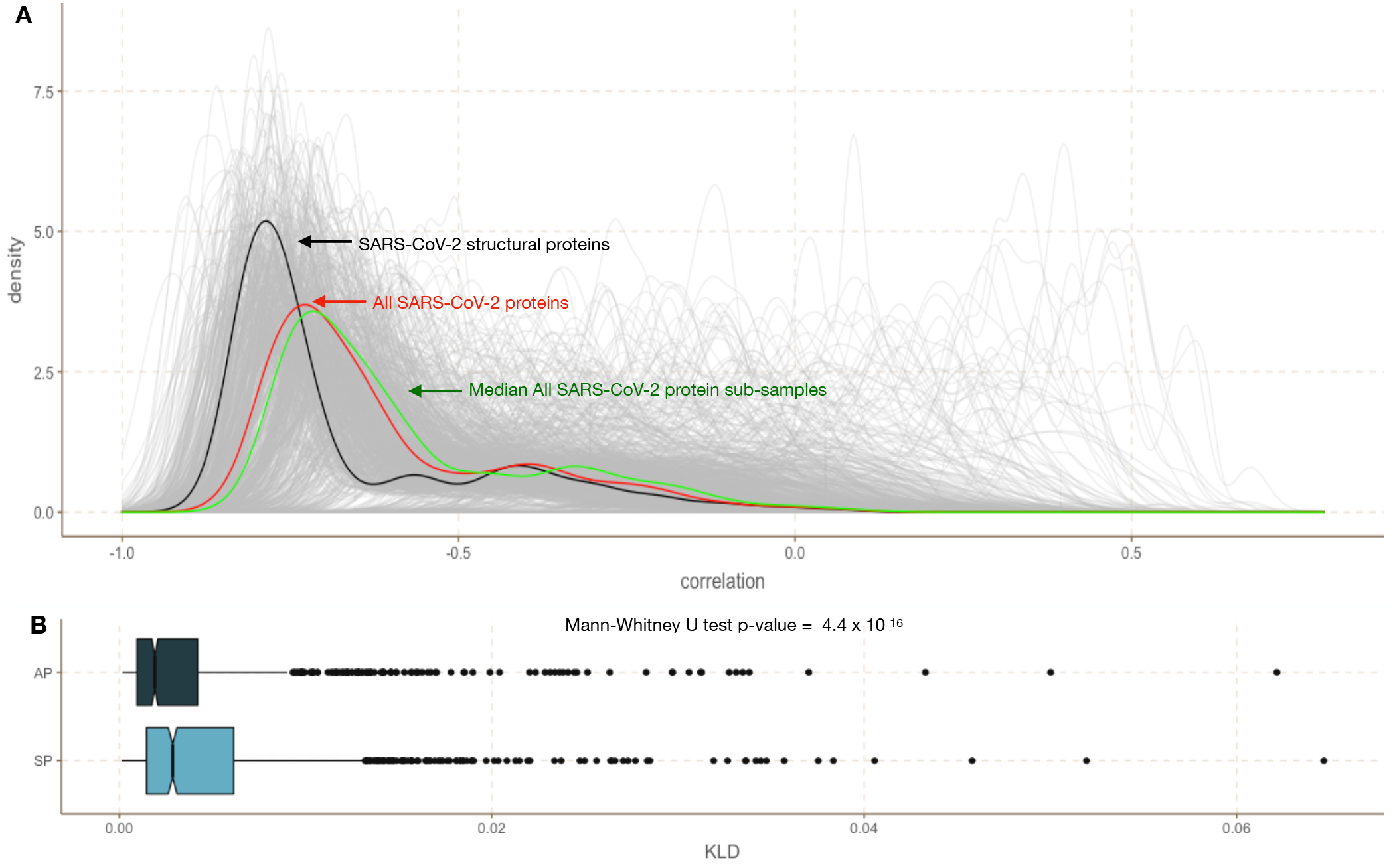
The effect of sub-sampling EnsembleMHC-identified MHC-I peptides derived from the full SARS-CoV-2 proteome on EMP score -deaths per million correlation distribution. The robustness of the observed distinction between the EMP score -deaths per million correlation distributions between SARS-CoV-2 structural proteins and all SARS-CoV-2 proteins was assessed by performing sub-sampling of non-structural SARS-CoV-2 MHC-I peptides. **A**, 1,000 sub-sampling iterations were performed by randomly selecting 108 peptides from the full SARS-CoV-2 proteome that passed the 5% *peptide*^*FDR*^ filter. The correlation between the population EMP score produced by each sub-sampled set of peptides and observed deaths per million were plotted (grey lines). The correlation distribution observed for identified SARS-CoV-2 structural protein peptides (black line), all SARS-CoV-2 proteins (red line), and the median correlation distribution across all subsampling iterations (green line) were plotted for comparison. **B**, Kullback-Leibler divergence was calculated for the correlation distribution of each down sample iteration relative to either the correlation distribution of the all peptide group (AP) or the structural peptide group (SP).

**Figure A.19:**
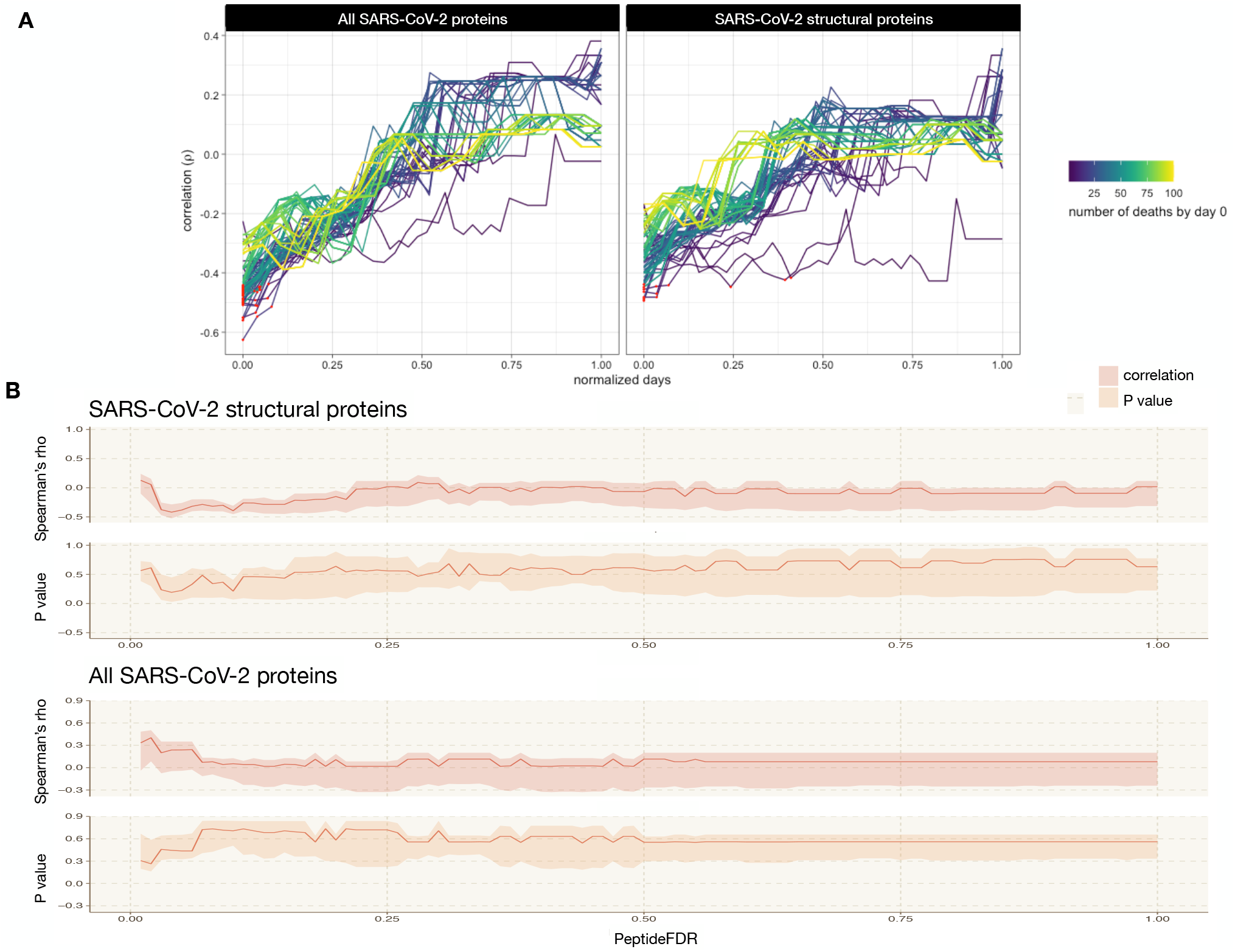
The effect of MHC shuffling on the correlation of EMP score and deaths per million. **A,** The MHC-I allele assessment of peptides that passed an individual algorithm binding affinity thresholds were shiffled prior to *peptide*^*FDR*^ filtering. The red points indicate correlations with a p-value ≤ 5%. **B**, The impact of varying *peptide*^*FDR*^ cutoff threshold on the shuffled MHC data set. For each *peptide*^*FDR*^ cutoff threshold (x-axis), the upper bound of the shaded region indicates the 75^*th*^ percentile, the lower bound indicates the 25^*th*^ percentile, and the solid line indicates the median.

**Figure A.20:**
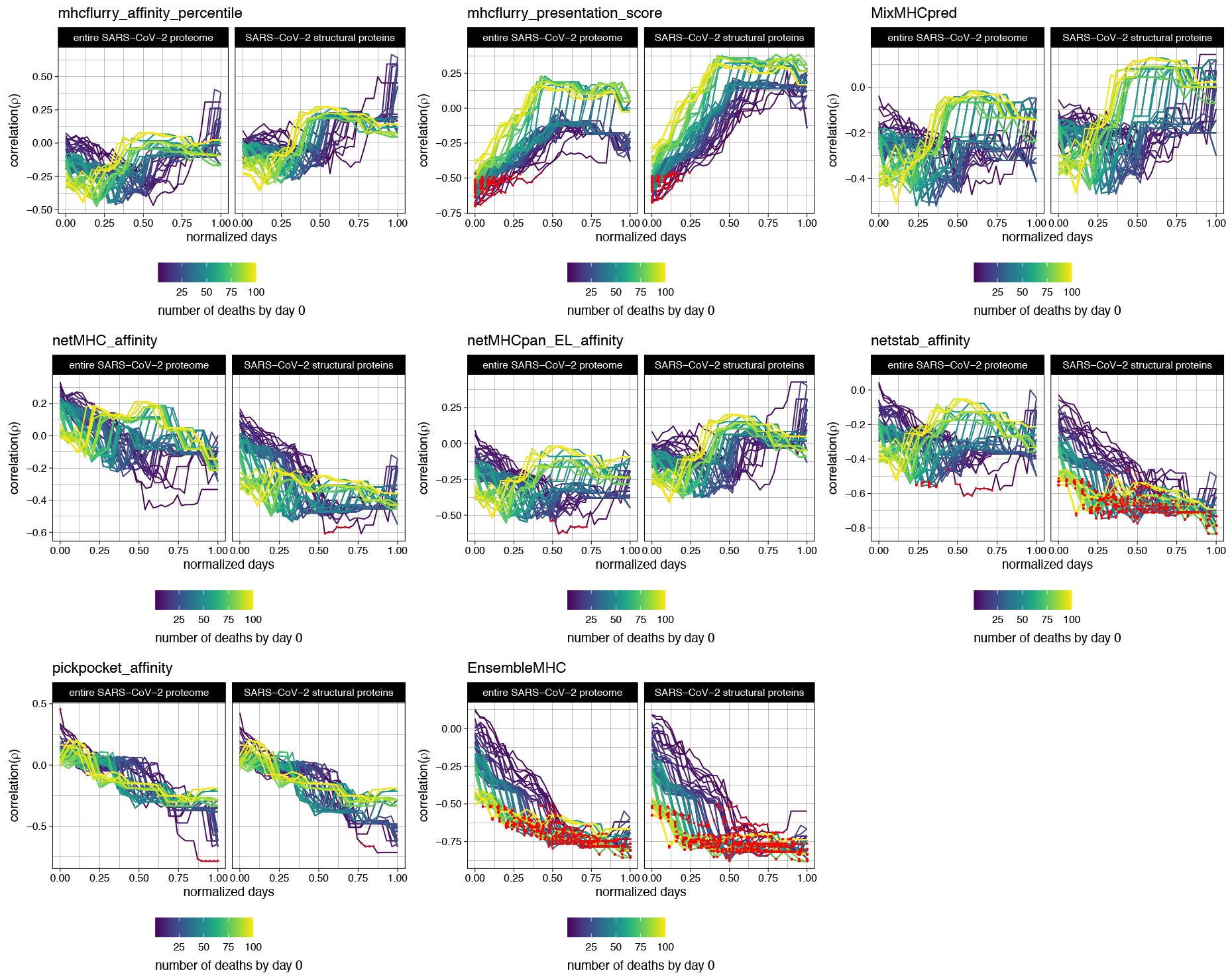
Individual algorithms are unable to fully recreate the correlation with population mortality reported by EnsembleMHC. Population SARS-CoV-2 binding capacities using only single algorithms were correlated to observed deaths per million. For each algorithm, the population SARS-CoV-2 binding capacity was calculated from the resulting viral peptide-MHC allele distribution using restrictive MHC-I binding affinity cutoffs (≤ 0.5% for binding percentile scores, top 0.5% MHCflurry presentation score, and ≤ 50*nm* for PickPocket). Red points indicate a PPV ≥ 95%.

**Figure A.21:**
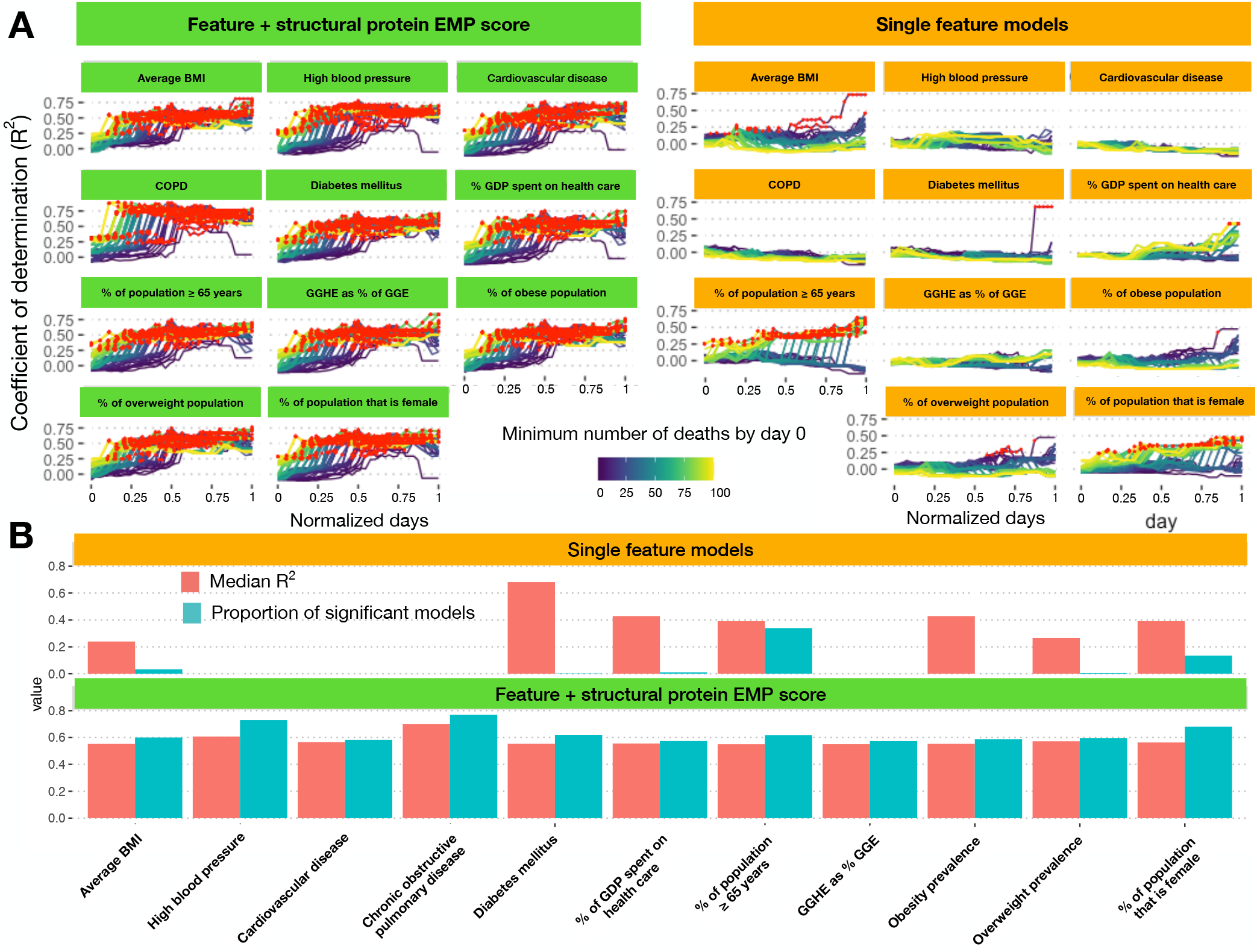
Addition of structural protein EMP score significantly improves linear model fit to observed deaths per million. **A,** Linear models were constructed using either a single risk factor (yellow) or a combination of a risk factor and structural protein EMP scores (green). The x-axis indicates the number of normalized days from when a minimum death threshold was met (line color), and the y-axis indicates the observed adjusted *R*^2^ value. **B**, A summary of results obtained from single feature linear models (top panel, yellow) or the combination models (bottom panel, green). The red bars indicate the median *R*^2^ value achieved by that model and the blue bars indicate the proportion of regressions that were found to be significant (F-test ≤ 0.05).

**Figure A.22:**
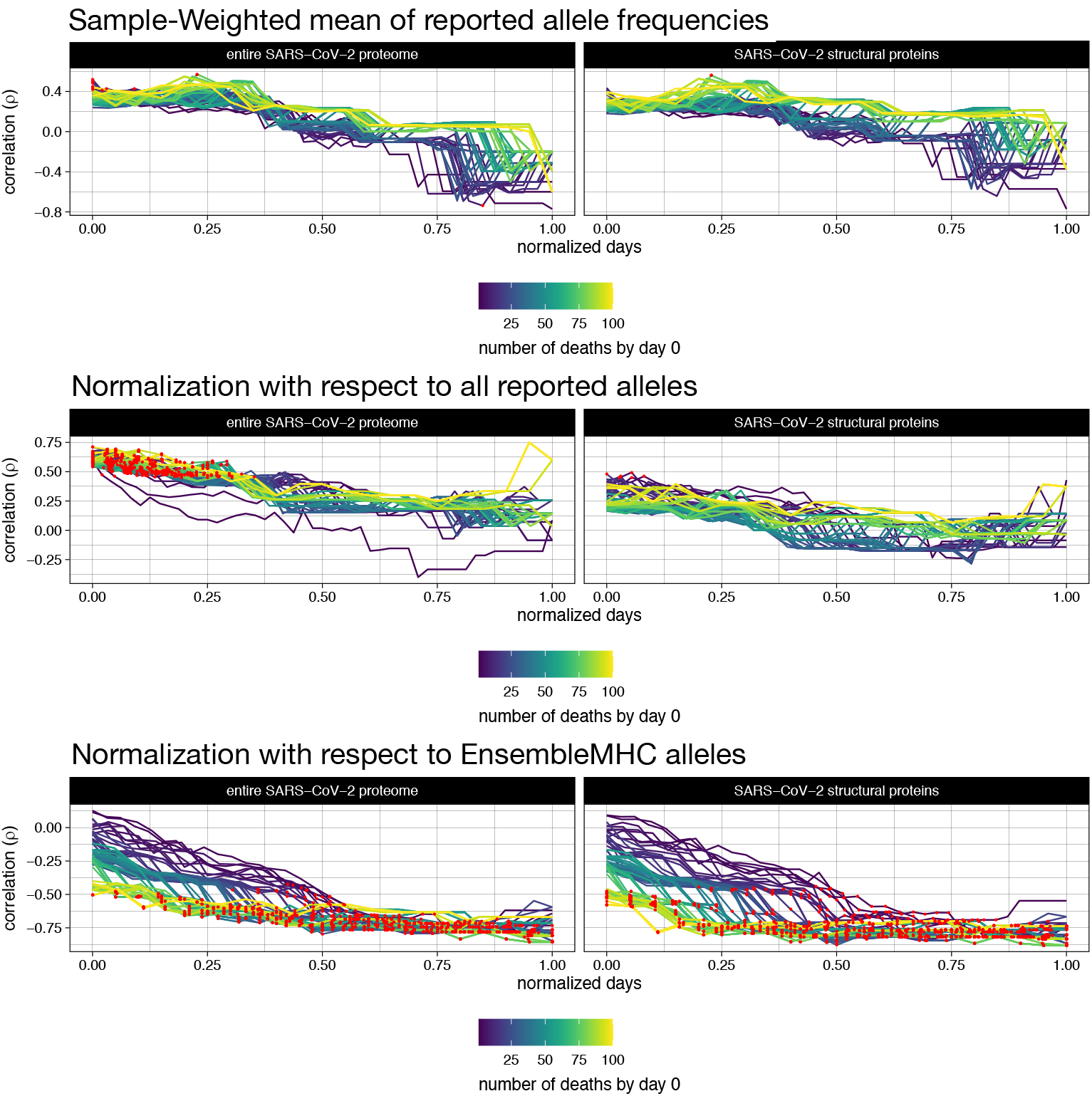
EnsembleMHC population score and deaths per million correlation using different allele frequency accounting methods. The effect of different allele frequency normalization techniques on the reported correlations between SARS-CoV-2 mortality and EMP scores based on the full SARS-CoV-2 proteome (left column) or SARS-CoV-2 structural proteins (right column). **Top panel**, The aggregation of allele frequencies within a particular country by taking the sample-weighted mean of reported frequencies for the 52 selected MHC-I alleles. **Middle panel**, Normalizing allele count with respect to all detected alleles in a given population. **Bottom panel**, Normalizing allele count with respect to only the 52 select alleles.

**Figure A.23:**
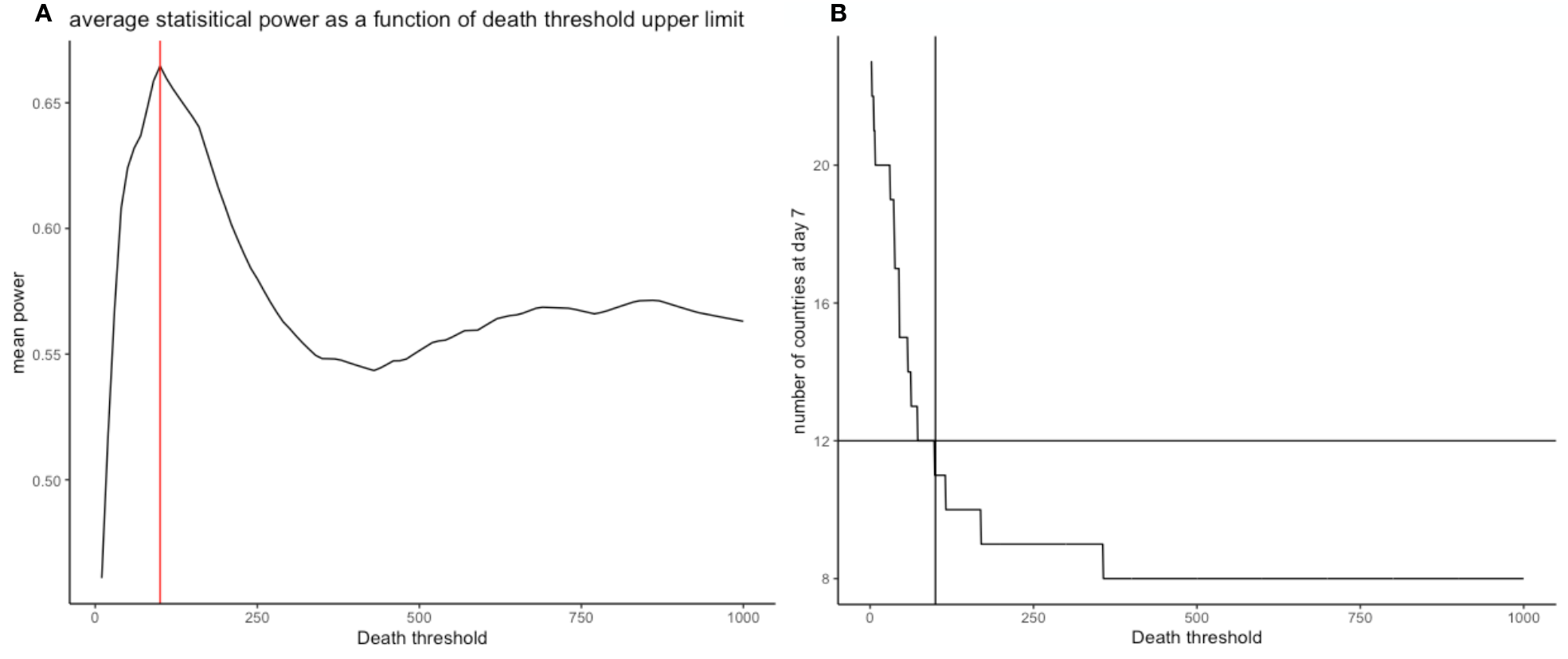
Justification of upper limit for death threshold. **A,** The mean statistical power of all resulting correlations between EnsembleMHC population scores and observed deaths per million at different minimum reported death thresholds. The red line indicates a minimum death threshold of 100 deaths by day 0, the selected upper limit for analysis. **B**, The number of countries remaining at day seven using different minimum death thresholds. The cross bar indicates that there would be more than half of the considered countries remaining at day 7 when using the 100 minimum death threshold.

**Figure A.24:**
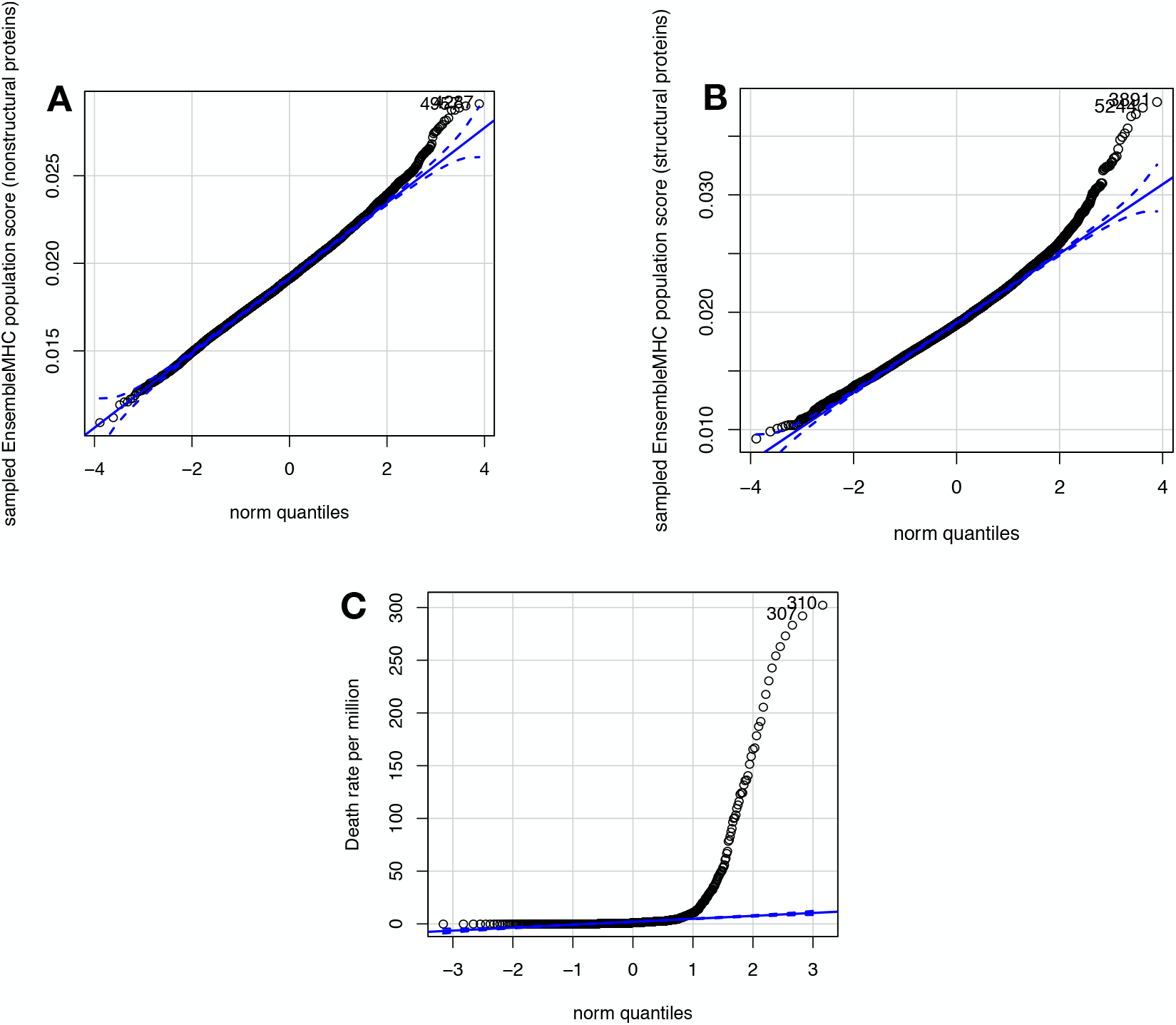
Justification of non-parametric correlation analysis. The use of non-parametric correlation analysis, namely spearman’s rho, is justified by the non-normality of the underlying data. EnsembleMHC population scores based on the full SARS-CoV-2 proteome and SARS-CoV-2 structural proteins were calculated for 10,000 simulated countries. Allele frequencies for simulated countries were generate by randomly sampling an observed allele frequency for each of the 52 alleles and re-normalizing to ensure the sum of allele frequencies were equal to one. **A**, The Q-Q plot for the simulated EnsembleMHC population score distribution based on the full SARS-CoV-2 proteome. **B**, The Q-Q plot for the simulated EnsembleMHC population score distribution based on SARS-CoV-2 structural proteins. **C**, The QQ plot for all reported deaths per Million. All three quantities show a considerable level of positive skewing, indicating non-normality.

**Figure A.25:**
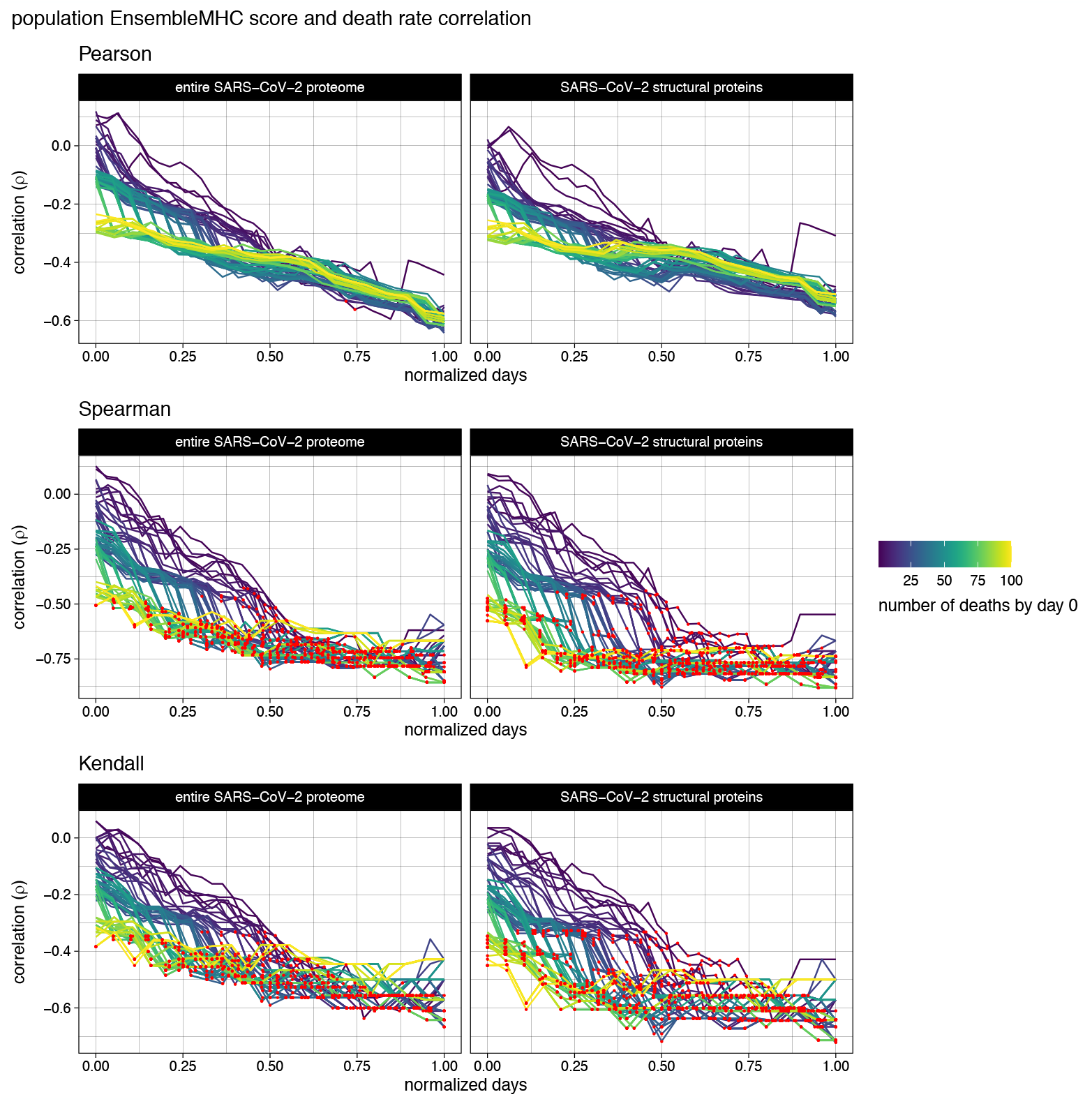
The effect of different correlation methods on the relationship between EnsembleMHC score and deaths per million. The correlation between EnsembleMHC population score with respect to all SARS-CoV-2 proteins **(left column)** or SARS-CoV-2 structural proteins **(right columns)** and deaths per million using Pearson’s r **(top)**, Spearman’s rho **(middle)**, and Kendall’s tau **(bottom)**. Correlations that were shown to be statistically significant are colored with a red point.

## B tables

**Figure B.1:** MHC-I peptide identified by Ensemble MHC. All identified peptides with a *peptide*^*FDR*^ ≤ 0.05 (not pictured due to size).

**Figure B.2:**
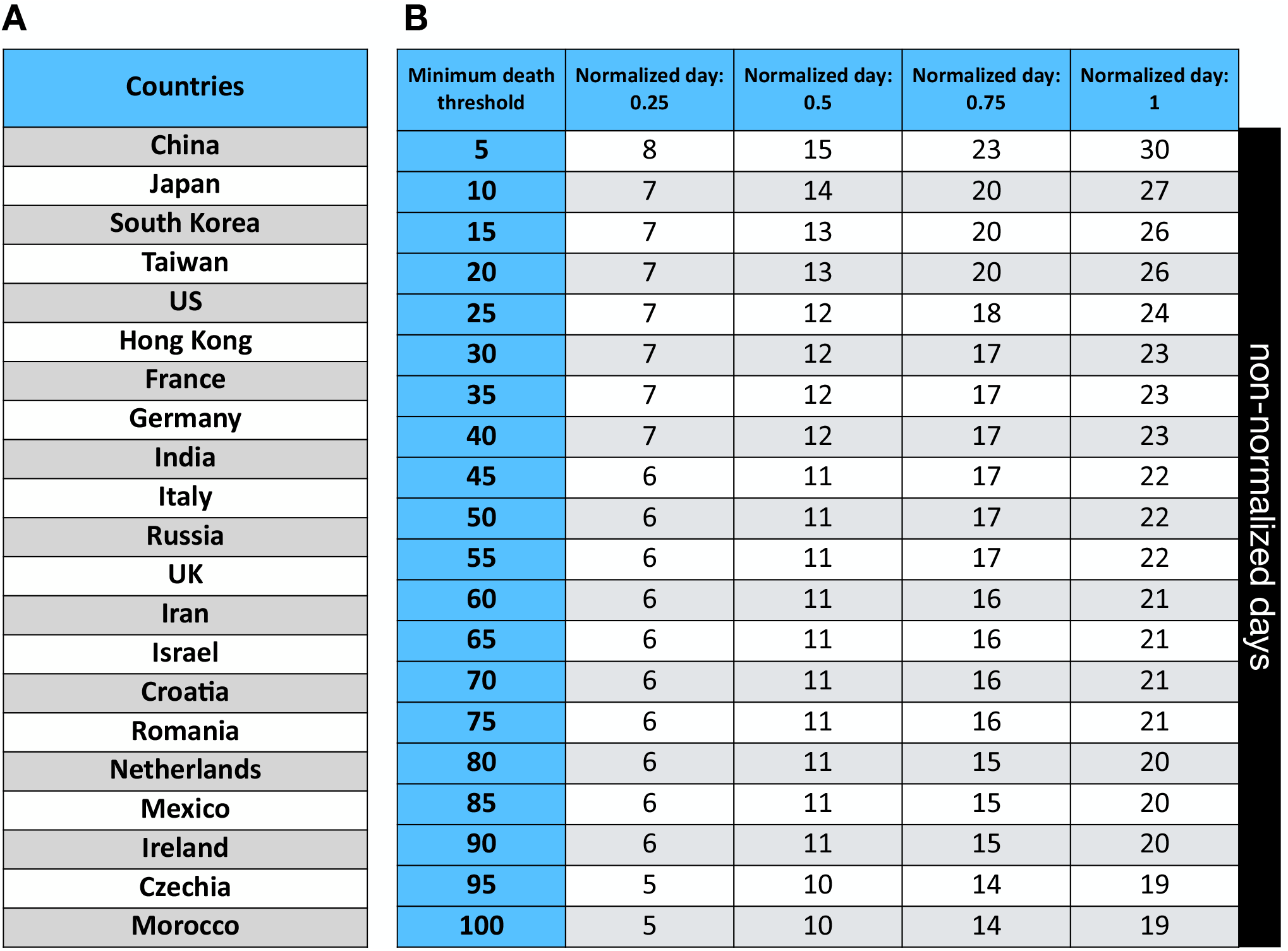
Description of the additional socioeconomic factors selected for analysis. **A,** The 23 countries for which SARS-CoV-2 population binding capacities were calculated. **B**, The mapping of normalized days to real days for normalized day quartiles (0.25,, 0.5, 0.75, 1) at select minimum death thresholds.

**Figure B.3:** EMP score correlation data. All correlation data pertaining to the correlations between EMP score and deaths per million. This includes rho estimate, 95% CI, non-normalized days, and sample size for each correlation (not pictured due to size).

**Figure B.4:**
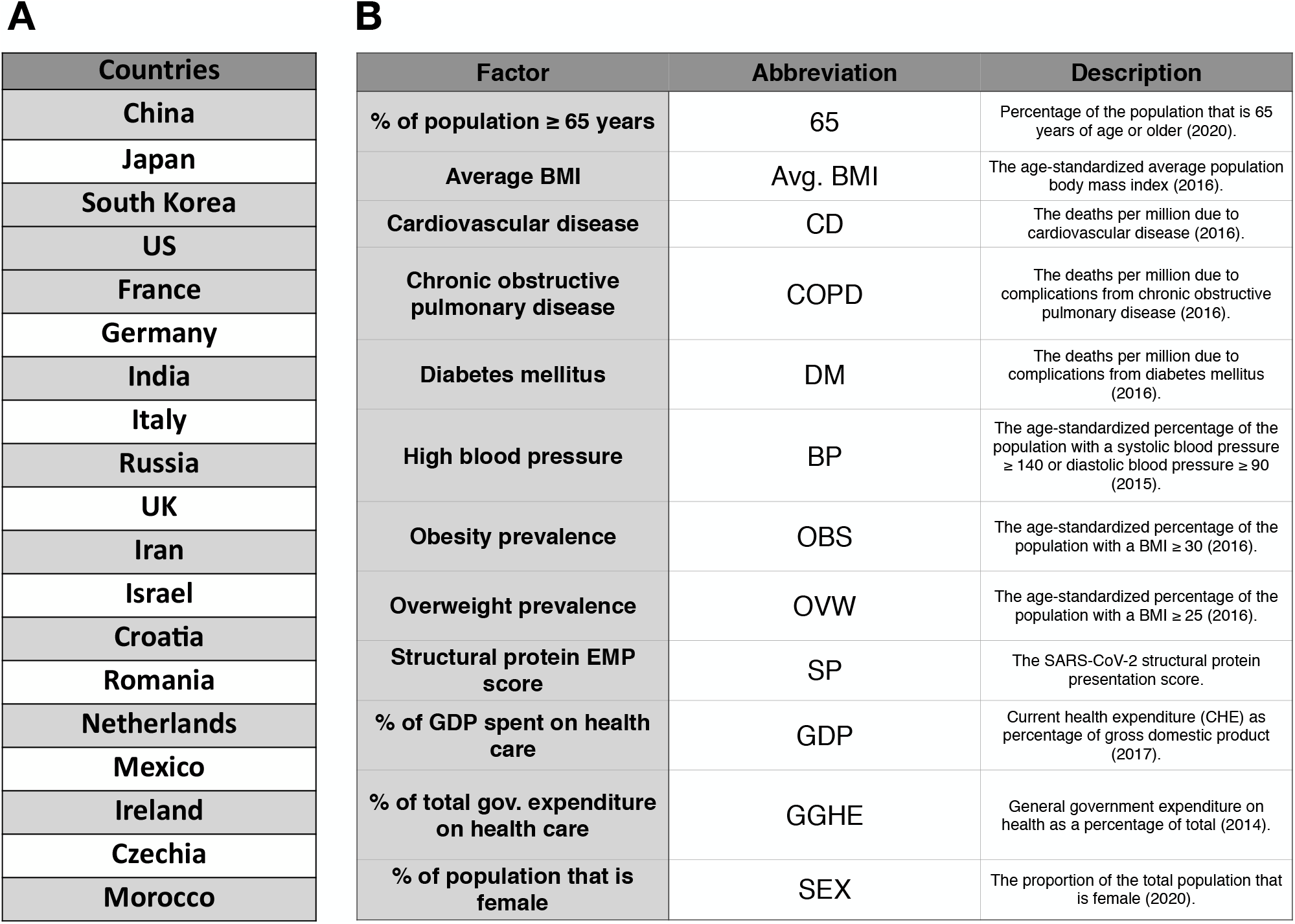
Socioeconomic and health-related risk factors. **A,** 21 countries were selected for analysis based on the existence of data in the Global Health Observatory data repository and inclusion in the 23 country set used for EMP score analysis. **B**, Descriptions and abbreviations for the selected risk factors. Each factor is labeled with the year that the data was collected. In every case, the most recent data was selected for analysis.

## Notes

### Competing Interest Statement

The authors have declared no competing interest.

### Clinical Trial

N/A

### Summary of Updates

Additional analysis and reformatted figures.

## References

[1] Zi Yue Zu et al. “Coronavirus disease 2019 (COVID-19): a perspective from China”. In: Radiology (2020), p. 200490.

[2] Qun Li et al. “Early transmission dynamics in Wuhan, China, of novel coronavirus–infected pneumonia”. In: New England Journal of Medicine (2020).

[3] Yan-Rong Guo et al. “The origin, transmission and clinical therapies on coronavirus disease 2019 (COVID-19) outbreak–an update on the status”. In: Military Medical Research 7.1 (2020), pp. 1–10.

[4] Rudragouda Channappanavar, Jincun Zhao, and Stanley Perlman. “T cell-mediated immune response to respiratory coronaviruses”. In: Immunologic research 59.1-3 (2014), pp. 118–128.

[5] Hsueh-Ling Janice Oh et al. “Understanding the T cell immune response in SARS coronavirus infection”. In: Emerging microbes & infections 1.1 (2012), pp. 1–6.

[6] Oi-Wing Ng et al. “Memory T cell responses targeting the SARS coronavirus persist up to 11 years post-infection”. In: Vaccine 34.17 (2016), pp. 2008–2014.

[7] Nina Le Bert et al. “SARS-CoV-2-specific T cell immunity in cases of COVID-19 and SARS, and uninfected controls”. In: Nature 584.7821 (2020), pp. 457–462.

[8] Alba Grifoni et al. “Targets of T cell responses to SARS-CoV-2 coronavirus in humans with COVID-19 disease and unexposed individuals”. In: Cell (2020).

[9] Vasiliki Matzaraki et al. “The MHC locus and genetic susceptibility to autoimmune and infectious diseases”. In: Genome biology 18.1 (2017), p. 76.

[10] Marie Lin et al. “Association of HLA class I with severe acute respiratory syndrome coronavirus infection”. In: BMC Medical Genetics 4.1 (2003), p. 9.

[11] Sheng-Fan Wang et al. “Human-leukocyte antigen class I Cw 1502 and class II DR 0301 genotypes are associated with resistance to severe acute respiratory syndrome (SARS) infection”. In: Viral immunology 24.5 (2011), pp. 421–426.

[12] Margaret HL Ng et al. “Association of human-leukocyte-antigen class I (B* 0703) and class II (DRB1* 0301) genotypes with susceptibility and resistance to the development of severe acute respiratory syndrome”. In: Journal of Infectious Diseases 190.3 (2004), pp. 515– 518.

[13] MH Ng et al. “Immunogenetics in SARS: a case-control study.” In: Hong Kong medical journal= Xianggang yi xue za zhi 16.5 Suppl 4 (2010), p. 29.

[14] Alicia Sanchez-Mazas. “HLA studies in the context of coronavirus outbreaks”. In: Swiss Medical Weekly 150.1516 (2020).

[15] Austin Nguyen et al. “Human leukocyte antigen susceptibility map for SARS-CoV-2”. In: Journal of Virology (2020).

[16] Weilong Zhao and Xinwei Sher. “Systematically benchmarking peptide-MHC binding predictors: From synthetic to naturally processed epitopes”. In: PLoS computational biology 14.11 (2018).

[17] Siranush Sarkizova et al. “A large peptidome dataset improves HLA class I epitope prediction across most of the human population”. In: Nature Biotechnology 38.2 (2020), pp. 199–209.

[18] Faviel F González-Galarza et al. “Allele frequency net 2015 update: new features for HLA epitopes, KIR and disease and HLA adverse drug reaction associations”. In: Nucleic acids research 43.D1 (2015), pp. D784–D788.

[19] Timothy J O’Donnell, Alex Rubinsteyn, and Uri Laserson. “MHCflurry 2.0: Improved Pan-Allele Prediction of MHC Class I-Presented Peptides by Incorporating Antigen Processing”. In: Cell Systems (2020).

[20] Vanessa Jurtz et al. “NetMHCpan-4.0: improved peptide– MHC class I interaction predictions integrating eluted ligand and peptide binding affinity data”. In: The Journal of Immunology 199.9 (2017), pp. 3360–3368.

[21] Massimo Andreatta and Morten Nielsen. “Gapped sequence alignment using artificial neural networks: application to the MHC class I system”. In: Bioinformatics 32.4 (2016), pp. 511–517.

[22] Michal Bassani-Sternberg et al. “Deciphering HLA-I motifs across HLA peptidomes improves neo-antigen predictions and identifies allostery regulating HLA specificity”. In: PLoS computational biology 13.8 (2017), e1005725.

[23] Hao Zhang, Ole Lund, and Morten Nielsen. “The Pick-Pocket method for predicting binding specificities for receptors based on receptor pocket similarities: application to MHC-peptide binding”. In: Bioinformatics 25.10 (2009), pp. 1293–1299.

[24] Michael Rasmussen et al. “Pan-specific prediction of peptide–MHC class I complex stability, a correlate of T cell immunogenicity”. In: The Journal of Immunology 197.4 (2016), pp. 1517–1524.

[25] Sinu Paul et al. “HLA class I alleles are associated with peptide-binding repertoires of different size, affinity, and immunogenicity”. In: The Journal of Immunology 191.12 (2013), pp. 5831–5839.

[26] K Nichols. “False discovery rate procedures”. In: Statistical Parametric Mapping. Elsevier, 2007, pp. 246– 252.

[27] Morten Nielsen et al. “Immunoinformatics: Predicting Peptide–MHC Binding”. In: Annual Review of Biomedical Data Science 3 (2020).

[28] Thomas Trolle et al. “The length distribution of class I– restricted T cell epitopes is determined by both peptide supply and MHC allele–specific binding preference”. In: The Journal of Immunology 196.4 (2016), pp. 1480– 1487.

[29] Nicolas Rapin et al. “The MHC motif viewer: a visualization tool for MHC binding motifs”. In: Current protocols in immunology 88.1 (2010), pp. 18–17.

[30] Yoav Benjamini and Yosef Hochberg. “Controlling the false discovery rate: a practical and powerful approach to multiple testing”. In: Journal of the Royal statistical society: series B (Methodological) 57.1 (1995), pp. 289– 300.

[31] Elizabeth J Williamson et al. “OpenSAFELY: factors associated with COVID-19 death in 17 million patients”. In: Nature (2020), pp. 1–11.

[32] Simon de Lusignan et al. “Risk factors for SARS-CoV-2 among patients in the Oxford Royal College of General Practitioners Research and Surveillance Centre primary care network: a cross-sectional study”. In: The Lancet Infectious Diseases (2020).

[33] Morgane Rolland et al. “Broad and Gag-biased HIV-1 epitope repertoires are associated with lower viral loads”. In: PloS one 3.1 (2008).

[34] Katie M Campbell et al. “Prediction of SARS-CoV-2 epitopes across 9360 HLA class I alleles”. In: bioRxiv (2020).

[35] Diego Chowell et al. “Evolutionary divergence of HLA class I genotype impacts efficacy of cancer immunotherapy”. In: Nature medicine 25.11 (2019), pp. 1715–1720.

[36] Jatin Arora et al. “HLA heterozygote advantage against HIV-1 is driven by quantitative and qualitative differences in HLA allele-specific peptide presentation”. In: Molecular biology and evolution 37.3 (2020), pp. 639– 650.

[37] Nathan P Croft et al. “Most viral peptides displayed by class I MHC on infected cells are immunogenic”. In: Proceedings of the National Academy of Sciences 116.8 (2019), pp. 3112–3117.

[38] Yanan Cao et al. “Comparative genetic analysis of the novel coronavirus (2019-nCoV/SARS-CoV-2) receptor ACE2 in different populations”. In: Cell discovery 6.1 (2020), pp. 1–4.

[39] Marek Prachar et al. “COVID-19 Vaccine Candidates: Prediction and Validation of 174 SARS-CoV-2 Epitopes”. In: bioRxiv (2020).

[40] Syed Faraz Ahmed, Ahmed A Quadeer, and Matthew R McKay. “COVIDep: A web-based platform for real-time reporting of vaccine target recommendations for SARS-CoV-2”. In: Nature reviews microbiology 15 (2020), pp. 2141–2142.

[41] Helen M Berman et al. “The protein data bank”. In: Nu-cleic acids research 28.1 (2000), pp. 235–242.

[42] Chengxin Zhang et al. “Protein structure and sequence re-analysis of 2019-nCoV genome refutes snakes as its intermediate host or the unique similarity between its spike protein insertions and HIV-1”. In: Journal of proteome research (2020).

[43] William Humphrey, Andrew Dalke, Klaus Schulten, et al. “VMD: visual molecular dynamics”. In: Journal of molecular graphics 14.1 (1996), pp. 33–38.

[44] Ensheng Dong, Hongru Du, and Lauren Gardner. “An interactive web-based dashboard to track COVID-19 in real time”. In: The Lancet infectious diseases (2020).

[45] Katherine S Button et al. “Power failure: why small sample size undermines the reliability of neuroscience”. In: Nature Reviews Neuroscience 14.5 (2013), pp. 365–376.

[46] Andrew P Ferretti et al. “COVID-19 patients form memory CD8+ T cells that recognize a small set of shared immunodominant epitopes in SARS-CoV-2”. In: (2020).

[47] Annika Nelde et al. “SARS-CoV-2-derived peptides define heterologous and COVID-19-induced T cell recognition”. In: Nature immunology (2020), pp. 1–12.

[48] Thomas M Snyder et al. “Magnitude and dynamics of the T-cell response to SARS-CoV-2 infection at both individual and population levels”. In: medRxiv (2020).

[49] Ahmed A Quadeer, Syed Faraz Ahmed, and Matthew R McKay. “Epitopes targeted by T cells in convalescent COVID-19 patients”. In: bioRxiv (2020).

